# Major depression and atherosclerotic disease: Linking shared genetics to pathways in blood, brain, heart, and atherosclerotic plaques

**DOI:** 10.1101/2025.07.11.25331336

**Authors:** Emma Pruin, Meike Bartels, Ernest Diez Benavente, Noortje A.M. van den Dungen, Joost K.R. Hoekstra, Dominique D.P. de Kleijn, Lennart P.L. Landsmeer, Michal Mokry, Gerard Pasterkamp, Brenda W.J.H. Penninx, Wouter J. Peyrot, Hester M. den Ruijter, Sander W. van der Laan, Yuri Milaneschi

## Abstract

**Background:** The increased risk of atherosclerotic diseases (stroke, coronary artery disease [CAD]) observed in depression may stem from shared pathophysiology. We examined whether: 1) major depression (MD) and atherosclerotic traits share genetic risk, and 2) altered gene expression in various tissues linked to shared genetics has a potential causal role in depression etiology.

**Methods:** Data from the largest genome-wide association studies of MD (N=3,887,532) and 8 atherosclerotic traits (N=26,909-1,308,460) were used in Mendelian randomization and colocalization to detect cross-trait causal associations and genomic loci containing shared causal variants. In shared loci, summary data-based Mendelian randomization estimated the effects of gene expression on MD etiology using expression quantitative trait loci datasets from whole blood, brain and heart tissues and atherosclerotic plaques from the Athero-Express Biobank Study.

**Results:** MD genetic liability increased risk of any stroke (OR=1.15, p=9.47×10^-8^), ischemic stroke (OR=1.16, p=1.52×10^-7^), small vessel disease (OR=1.34, p=4.76×10^-5^) and CAD (OR=1.2, 95%CIs=1.13-1.26, p=3.76×10^-22^). Eight genomic regions harbored potentially shared causal variants, including one on chromosome 7 linking MD with any stroke, ischemic stroke and CAD. Altered expression of 16 genes in blood, 10 in brain, and 6 in heart was found causal for MD etiology. In atherosclerotic plaques, one gene was linked to MD at nominal significance only.

**Conclusion:** Major depression and atherosclerotic diseases share genetic risk potentially acting in depression pathophysiology through expression of genes in blood, brain and heart tissues. An involvement of atherosclerotic plaques in depression etiology was not supported. Identified pathways could guide the development of new treatments to prevent depression-heightened atherosclerotic risk.

## Introduction

Depression is a highly prevalent disorder – about 1 in 6 people are affected at some point of their lives^1^ – and ranks amongst the top contributors to disability worldwide^2^. Its health burden is increased by high comorbidity with somatic disease^3^, such as an almost two-fold increase in risk of atherosclerotic diseases^4^. The comorbidity of depression and atherosclerotic disease may be promoted and maintained by numerous factors. Besides shared sociodemographic, behavioral and health-related (e.g. medication effects) determinants, biological pathways including inflammation and metabolic alterations could play an important role^5^. Aspects of this shared biology may represent promising targets for treatments to reduce the health impact of depression.

Consistent with the hypothesis of shared biological pathways, the comorbidity of depression and atherosclerotic disease has a substantial genetic basis^6^. The two conditions also show overlap in the genetic factors that influence them, with significant genetic correlations found for major depression (MD) with coronary artery disease (CAD), as well as stroke^7^. In addition, there is evidence for a potential causal effect of depression genetic liability on atherosclerotic diseases (not *vice versa) from Mend*elian Randomization (MR) studies^8,9^. We can leverage such information on the shared genetic architecture to shed light on downstream biological pathways active in different tissues.

Gene expression, the transcription of DNA to RNA, leads to cell differentiation and in consequence differs between tissues in health and disease. Tissue-specific effects of genetic loci can be assessed by analyzing their effects on gene expression; such genetic variants are known as expression quantitative loci (eQTL). For instance, risk loci for MD have been linked to expression in certain brain tissues, such as the prefrontal cortex, anterior cingulate cortex and hippocampus, as well as in blood and adrenal glands^10^. As an intermediary step in the link between genes and traits^11^, gene expression data provide the opportunity to link risk variants for the comorbidity of depression and atherosclerotic disease to the pathology of culprit tissues.

Considering the emerging role of inflammation and metabolic alterations in depression, atherosclerotic plaque emerges as a tissue that could be proposed as a substantial source of biological dysregulations in depression pathophysiology. Atherosclerotic plaques consist of lipid deposits accompanied by an inflammatory response within the vascular wall^12^. Excess presence of atherosclerotic plaque is the hallmark of diseases like CAD and cerebrovascular disease, which can lead to stroke. Previous studies have not investigated whether altered gene expression in atherosclerotic plaques might be contributing to depression.

In the present study, we maximized GWAS sample size by using summary statistics of the most recent meta-analyses to date for MD^10^, CAD^13^, and stroke (subtypes)^14^. We combined complementary evidence from the novel approaches of two-sample Mendelian Randomization and colocalization to reliably identify regions likely to contain shared causal variants. Furthermore, we tested whether the expression in various relevant tissues (brain regions, parts of the heart, and whole blood) of genes localized in shared loci was causally involved in depression etiology. Finally, leveraging unique eQTL data of atherosclerotic plaques from the Athero-Express Biobank Study^15^ - we investigated the potential causal links between gene expression in atherosclerotic plaques and depression risk.

## Methods and Materials

### GWAS summary statistics

We identified the largest meta-analysis of GWAS available to date of MD, a broad operationalization of depression combining different measures from clinical/interview, electronic health records, questionnaires, or self-report of diagnosis (N = 3,887,532), as well as eight atherosclerotic traits (stroke, ischemic stroke, small vessel disease, cardioembolic stroke, CAD, coronary artery calcification, carotid intima media thickness; N = 26,909 - 1,308,460), and obtained the respective summary statistics for European ancestry individuals (**Supplemental Table 1**).

To globally quantify the degree of genetic overlap between MD and the atherosclerotic traits we estimated single-nucleotide polymorphism (SNP) based genetic correlations using the related GWAS summary statistics and the high-definition likelihood (HDL) method^16^.

### Study design

Figure 1 illustrates the study design based on a sequence of interdependent analyses, with results from Mendelian Randomization and colocalization informing the subsequent analysis of various eQTL datasets. All analyses were based on the summary statistics of the selected GWAS.

**Figure 1.**
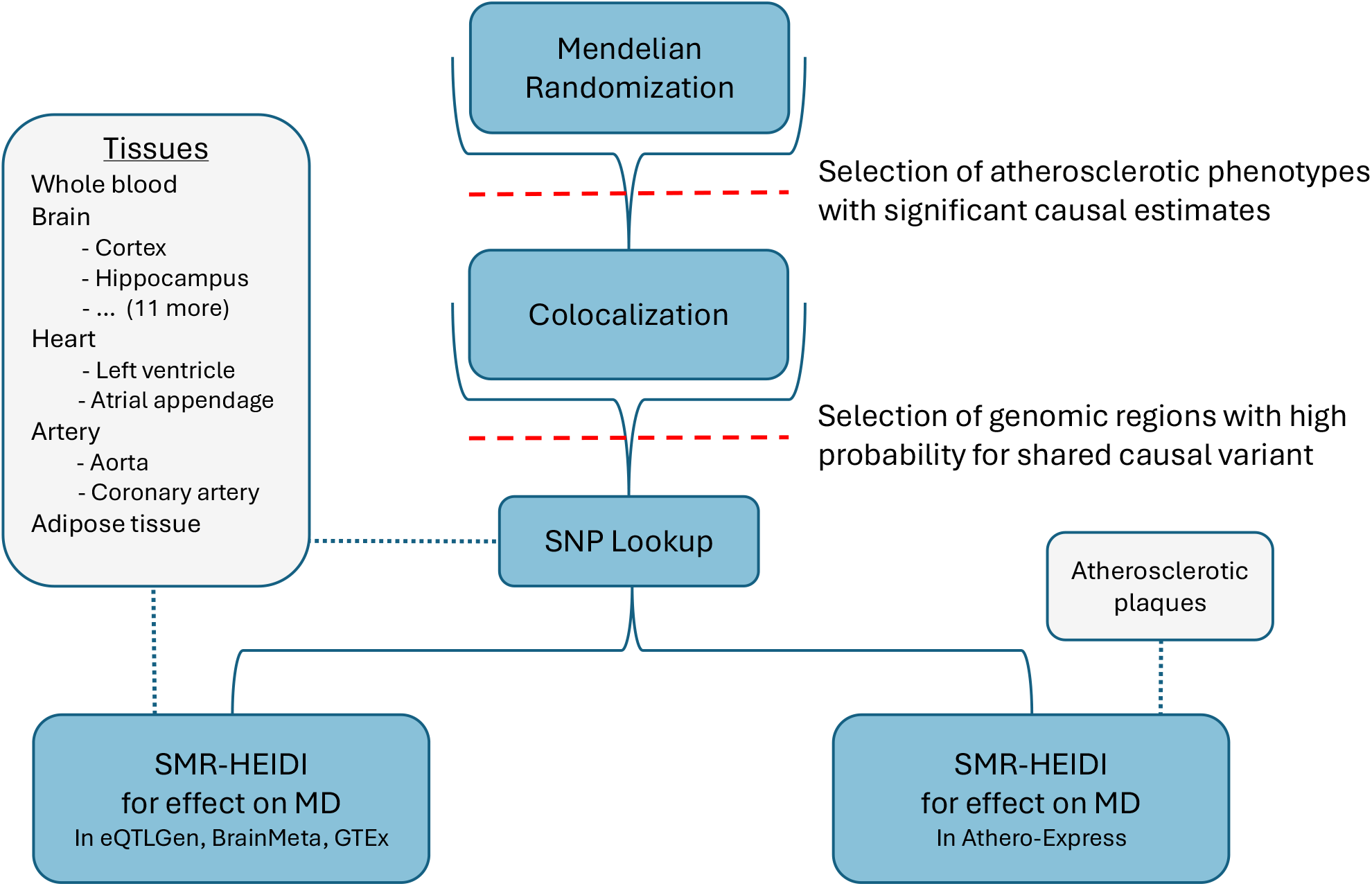
Flow of analyses. *SMR-HEIDI*: summary data–based Mendelian randomization and heterogeneity in dependent instruments method, *MD*: major depression, *eQTLGen/BrainMeta/GTEx/Athero-Express*: studies with eQTL data used here.

### Causal inference through Mendelian Randomization

We conducted Mendelian Randomization analyses using the R package TwoSampleMR (version 0.6.4)^17^. The instrumental variable consisted of several independent SNPs, selected from the exposure data by filtering and clumping (see **Supplementary Methods**). Causal estimates were obtained using the inverse variance weighted (IVW) method, which combines the ratio estimates of individual SNPs whilst inversely weighting them by their standard error^18^. Estimates were deemed significant when p < 0.1 after FDR correction for multiple testing (Benjamini-Hochberg procedure)^19^. Standard MR sensitivity analyses, described in the Supplementary Methods, were conducted for those trait pairs with significant causal estimates. Only traits with statistically significant MR estimates were selected for the next analysis steps (see **Figure 1**).

### Colocalization

Regions containing causal variants shared between MD and all atherosclerotic traits with a significant MR estimate were identified using Bayesian testing, as implemented by the `coloc` method^20^. First, regions of 1 megabase around the lead SNPs, selected for being significantly associated with MD^10^, were extracted from each of the GWAS datasets. The resulting pairs of summary statistics segments were analyzed using the R package coloc (version 5.2.3)^20^. Prior probabilities for SNP-trait associations were set to default (association with trait 1: p_1_ = 1×10^-4^, association with trait 2: p _2_= 1×10^-4^, association with both traits: p_12_ = 1×10^-5^). `Coloc` compares the following five hypotheses: The selected region contains no causal variant (H0), a causal variant for one of the traits (H1 and H2), non-shared causal variants for both traits (H3), or one shared causal variant for both traits (H4). We considered a posterior probability for H4 > 0.8 indicative of colocalization.

### SNP-Lookup in gene-expression databases

For each lead SNPs of a colocalizing region, we checked whether it was significantly associated with cross-tissue average expression of certain genes in the Genotype-Tissue Expression portal (https://www.gtexportal.org). We examined the eQTL’s tissue specificity based on its m-value^21^, a priori deeming whole blood, brain, heart, artery, and adipose tissue relevant for depression or atherosclerotic disease.

Additionally, we investigated the cellular expression of the genes previously identified as eQTL of the selected genetic variant in atherosclerotic disease. To this end, we used PlaqView (https://www.plaqview.com)^22^ which contains data from carotid atherosclerotic plaques from the Athero-Express Biobank Study (the Slenders et al. dataset^23^).

### SMR-HEIDI for effect on MD

The potential causal link of shared genomic regions with MD via gene expression was tested using summary data-based Mendelian randomization (SMR) and heterogeneity in dependent instruments (HEIDI) analysis^24^. SMR, a version of MR that was specifically developed to test the involvement of gene expression in disease mechanisms using eQTL data, is complemented by a HEIDI test, which rejects the hypothesis of shared causality or pleiotropy where linkage is a more likely explanation of an association. Here, we focused exclusively on regions of high likelihood for a shared causal variant between MD and atherosclerotic traits, as identified using colocalization analysis. We performed two main sets of SMR-HEIDI analyses across different tissues and eQTL datasets.

#### 1. Blood, brain and heart eQTL in publicly available datasets

First, we tested potential causal effects of gene expression in 20 different tissues on MD, by integrating the MD GWAS with publicly available eQTL data sets. These included eQTLGen (N = 31,684) for whole blood^25^, BrainMeta (N = 2,865) for brain tissue^26^, and GTEx for specific brain regions (N = 139 to 255), coronary artery (N = 240), aorta (N = 432), atrial appendage (N = 429) and left ventricle (N = 432)^27^, further details in **Supplemental Table 2**.

In the SMR-Portal^28^, p-value thresholds were set at 5.0×10^-8^ for the lead eQTL SNP and 1.57×10^-3^ for SNPs to be included in the HEIDI test. Estimates were deemed significant when p < 0.1 after FDR correction for multiple testing across eQTL-tissue pairs (Benjamini-Hochberg procedure)^19^.

#### 2. Atherosclerotic plaques in the Athero-Express Biobank study

The Athero-Express Biobank Study (AE) is an ongoing longitudinal biobank study including patients that undergo arterial endarterectomy in two Dutch tertiary referral centers since 2002^15^. For the present study, subsequent patients were included who underwent an endarterectomy of either the carotid or the femoral and iliac arteries and of which genotyping and transcriptomic data were available. Clinical data were extracted from patient medical files and standardized questionnaires. This study complies with the Declaration of Helsinki, and all participants provided informed consent. The medical ethical committees of the respective hospitals approved this study which was registered under number 22/018. Detailed description of the study procedures and generation of the eQTL datasets are provided in the **Supplemental Methods**. After quality control, eQTL data from 626 plaque samples were included.

Analysis of the plaque tissue differed from the analysis of the other tissues in being tailored to a dataset with overall weak associations. For this, we used the downloaded SMR software (version 1.3.1)^24^, allowing instruments with eQTL p-value < 0.1 after FDR correction for SNPs across pre-selected regions. The same procedure was repeated using CAD, stroke, and CIMT as outcomes, to better understand the role of eQTL in plaque pathology and disease risk.

## Results

We included GWAS summary statistics for MD and eight atherosclerotic traits (**Supplemental Table 1**). To explore whether these traits share common genetic pathways, suggestive of a potential causal relation, we calculated genetic correlations. Small genetic correlations (**Supplemental Table 3**) were detected of MD with any type of stroke (r=0.19, se=0.03, p=1.2×10^-12^), ischemic stroke (r=0.2, se=0.03, p=4.71×10^-13^), small vessel disease (r=0.24, se=0.05, p=7.4×10^-6^) and CAD (r=0.23, se=0.02, p=6.0×10^-32^).

### Genetic liability for major depression causally affects (ischemic) stroke and CAD

Next, we tested for a potential causal effect of MD liability on atherosclerotic traits, and vice versa. The strength of the MR instrumental variable, i.e. the effect of selected SNPs on the exposure, was found to be of adequate strength across traits (F > 10, see **Supplemental Table 5**). **Figure 2** shows that the genetic liability of MD significantly associated with an increased risk of any type of stroke (OR=1.15, 95%CIs=1.09-1.21, p=9.47×10^-8^), ischemic stroke (OR=1.16, 95%CIs=1.10-1.22, p=1.52×10^-7^), small vessel disease (OR=1.34, 95%CIs=1.16-1.55, p=4.76×10^-5^), and CAD (OR=1.2, 95%CIs=1.13-1.26, p=3.76×10^-22^), indicating a causal effect of MD on these diseases. The effects of MD genetic liability on cardioembolic stroke, large artery stroke, coronary artery calcification, and carotid intima media thickness, were not statistically significant. Causal estimates were robust, with the weighted median and MR Egger estimators showing the same direction and magnitude of effect as the IVW method.

**Figure 2.**
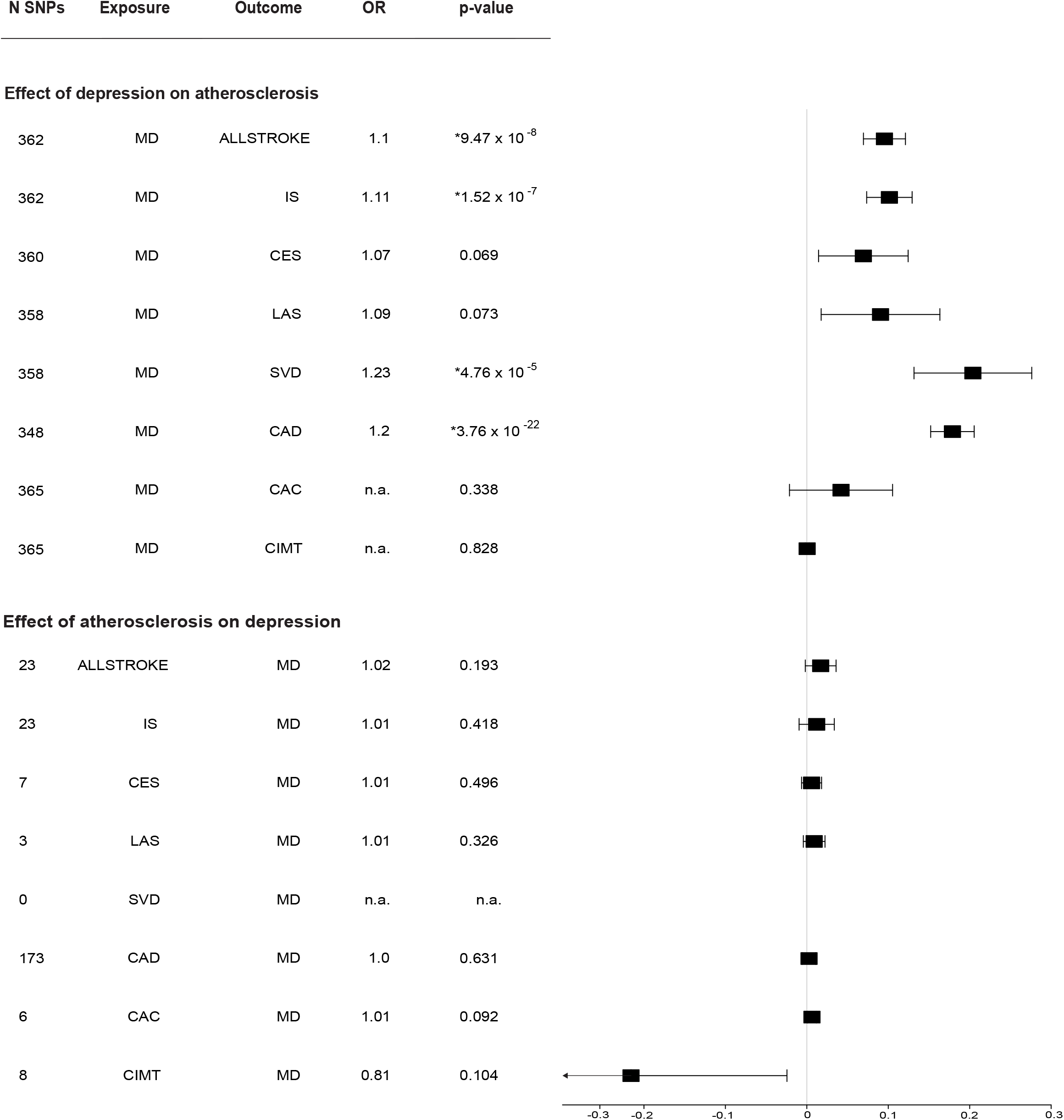
MR Analysis of Causal Effects between MD and Atherosclerosis. Forest plot displaying causal estimates (betas) from the IVW method. Error bars represent standard errors. * Indicates p-value significant at False Discovery Rate < 0.1. *N SNPs*: number of SNPs used as instruments for the exposure. *OR*: Odds ratio per doubling (2-fold increase) in the prevalence of the exposure (only for dichotomous outcomes). *ALLSTROKE*: any stroke, *IS*: ischemic stroke, *CES*: cardioembolic strokes, *LAS*: large artery stroke, *SVD*: small vessel disease, *CAD*: coronary artery disease, *CAC*: coronary artery calcification, *CIMT*: carotid intima-media thickness, *DEP*: major depression. *n*.*a*.: not applicable. Note that SVD was not included as an exposure, as no valid instruments were identified (N SNPs = 0). For full results refer to Supplemental Table 4 and 9.

For the results with significant effects – outcomes of stroke, ischemic stroke, small vessel disease, CAD – several sensitivity analyses were considered to assess the extent to which MR assumptions may have been violated (**Supplemental Tables 6-8**). Across traits, the Egger intercept did not indicate horizontal pleiotropy (not significantly different from 0), but Cochran’s Q statistic suggested instruments’ effects were heterogeneous (Q > 0, p < 0.05). MR PRESSO, which investigates pleiotropy by correcting for outlier SNPs, indicated horizontal pleiotropy for CAD (effect no longer significant after outlier detection and removal), but not for stroke and its subtypes.

In reversed MR analyses, the genetic liability of atherosclerotic diseases, as well as greater genetically predicted CAC and CIMT, were not significantly associated with increased risk of MD (see **Figure 2**), thus providing no evidence for a causal effect of atherosclerotic disease on depression.

### Major depression and atherosclerotic diseases share causal genetic loci

Given the expectation that traits with causal relationships found in MR are more likely to share causal genes, (ischemic) stroke, small vessel disease, and CAD were submitted to colocalization analysis with MD. A high probability of colocalization, with PP.H4 > 0.8, was found for MD and stroke at 2 lead SNPs, ischemic stroke at 1 lead SNPs and CAD at 7 lead SNPs (see **Supplemental Table 10** and **Supplemental Figures 1-10**). At an intron variant of the gene *TMEM106B on chromoso*me 7, rs10950392, there was a signal for colocalization of MD and all three atherosclerotic traits, see **Figure 3**. In addition, the region around this SNP showed the overall greatest probability of colocalization (PP.H4 = 99.82% for MD and CAD). No shared loci for MD and small vessel disease were found. Overall, these results provide evidence that regions surrounding eight unique SNPs are linked to potentially mechanistic pathways shared between depression and atherosclerotic disease.

**Figure 3.**
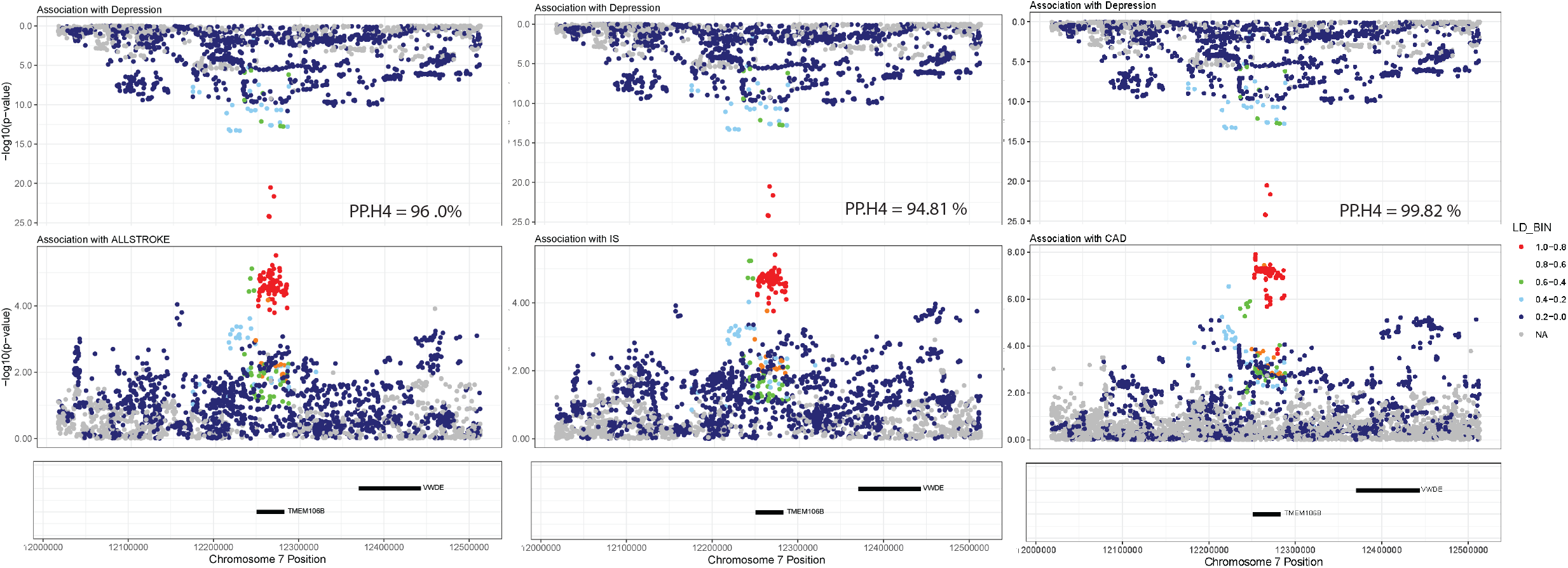
Mirror plots for top results of colocalization analysis. Three mirror plots of the region around lead SNP rs10950392. Panels from top to bottom: SNP associations with MD, SNP associations with the respective atherosclerotic disease, basic gene annotation. Plots were generated using the R package RACER (https://github.com/oliviasabik/RACER)^47^ . *ALLSTROKE: any stroke*; *IS*: ischemic stroke; *CAD*: coronary artery disease; *PP*.*H4*: posterior probability of coloc hypothesis for, i.e. the region containing a variant causal for both traits; *LD_BIN*: levels of linkage disequilibrium with rs10950392.

### Shared causal loci are associated with gene expression

GTEx lookup of lead SNPs found associations with the expression of twelve genes (see **Supplemental Figures 11-22**). For instance, the recurring lead SNP rs10950392, was found to be associated with expression *TMEM106B across tiss*ues, with highly tissue-specific expression relevant to depression and/or atherosclerosis, for instance in whole blood, left ventricle, atrial appendage and multiple brain regions.

### Shared causal loci act on major depression via tissue-specific gene expression

To investigate whether gene expression is involved in the pathway between shared genetic regions and MD, we conducted SMR analyses of eQTL in various tissues. The SMR analysis identified potential causal effects of gene expression on MD in all tested tissues, except for coronary artery and substantia nigra, see **Figure 4**. The greatest number of significant estimates was found for expression in blood, with seven genes (*ZFAND2A, TTYH3, PIP5K1C, GNA12, C7orf50, ARRDC5, ARL4A*) also passing the HEIDI test for linkage. In meta-analyzed data from all brain regions, expression of five genes (*RASGRP1, HDGFL2, GPR85, CHST12, AMZ1*) showed significant causal effects on MD. The expression of one gene, *MRM2*, showed significant causal effects on MD in all included GTEx tissues, with most of them also passing the HEIDI test. Notably, expression of *ZFAND2A* and *GNA12* in both blood and the heart (atrial appendage and left ventricle) had a causal effect on MD. Similarly, the effect of *PIP5K1C* was relevant across tissue domains, in whole blood, cerebellum and aorta. Besides, there was a positive relationship between number of significant estimates and sample size of eQTL datasets per tissue (**Supplemental Table 2**).

**Figure 4.**
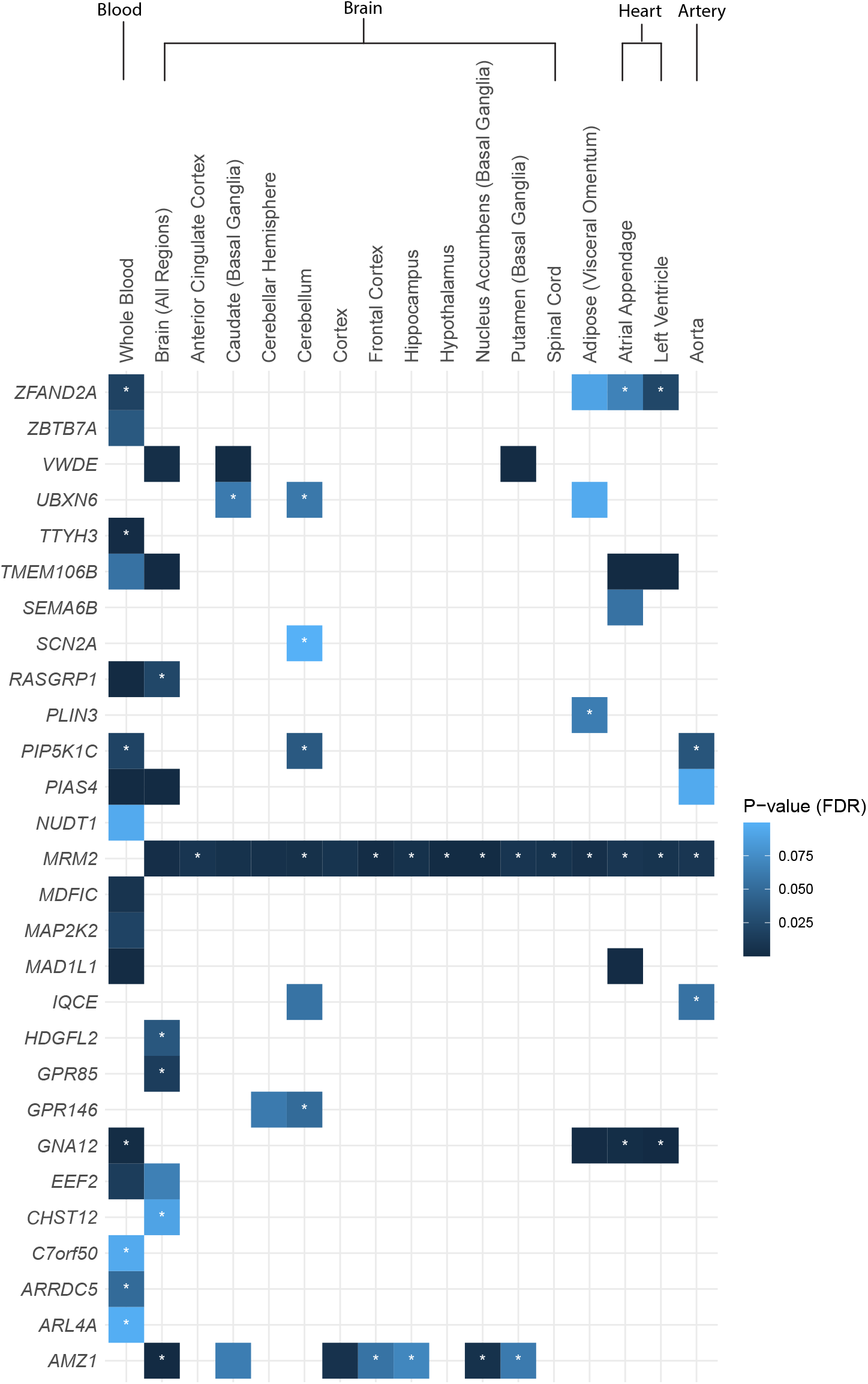
Significant causal estimates of gene expression in various tissues affecting MD. Plot mapping genes with significant causal estimates at FDR < 0.1 to the tissues where eQTL effects were found. * Indicates HEIDI p-value > 0.05, signifying that null hypothesis of a shared causal variant was not rejected. *FDR*: False discovery rate.

### Gene expression in atherosclerotic plaques was not linked to major depression

Finally, we investigated whether gene expression specific to atherosclerotic plaque tissue is causally linked to MD, by conducting SMR on eQTL data from 626 plaque samples. Samples stem from persons undergoing carotid endarterectomy, a cohort of older adults with high levels of disease and corresponding risk factors such as high levels of smoking (see **Table 1**) that is representative of the Athero-Express.

**Table 1.**
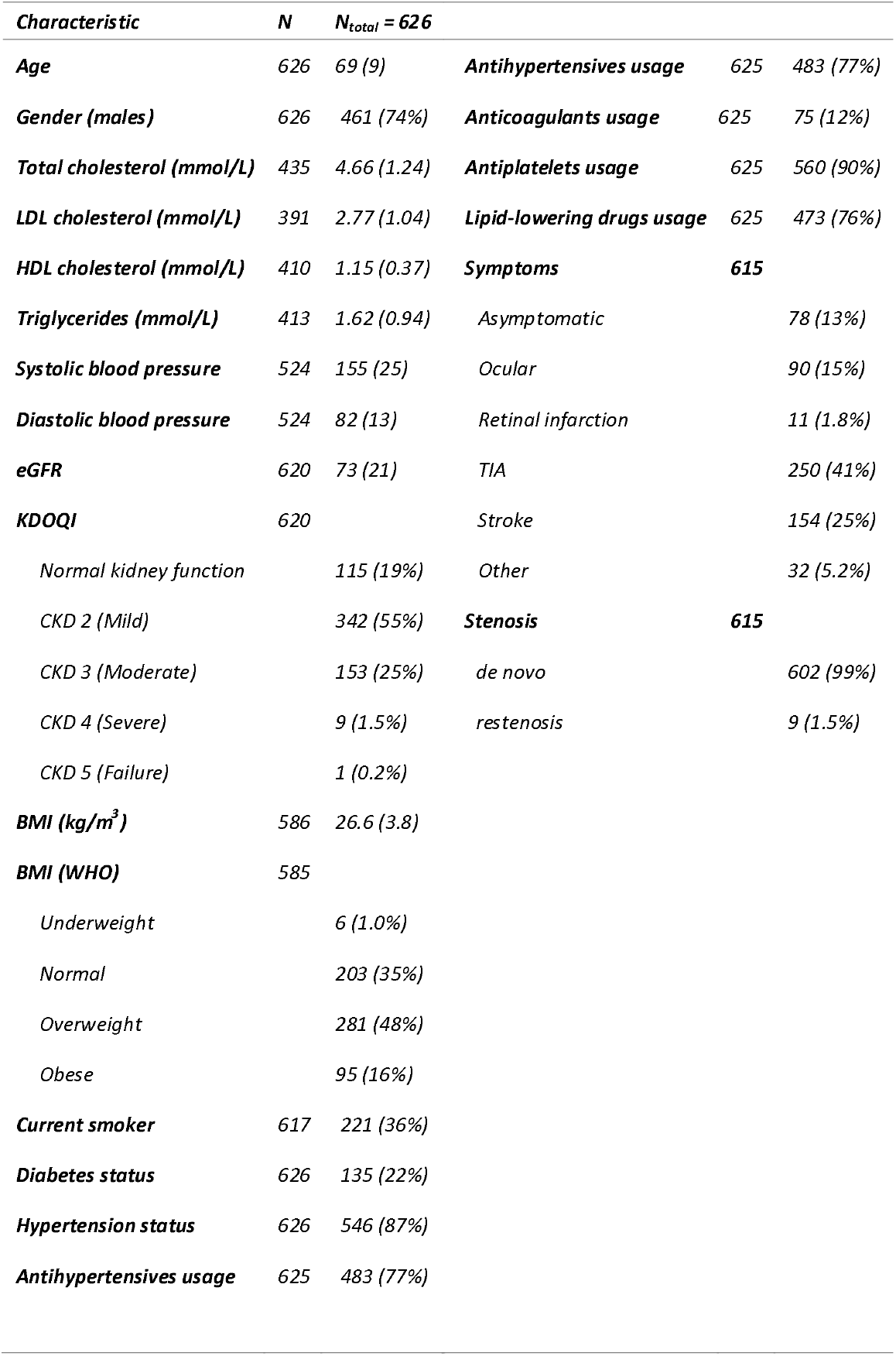
Baseline characteristics of the Athero-Express patients included in the eQTL analyses. *Characteristics* are given as n (%) or mean (standard deviation); *eGFR*: estimated glomerular filtration based on Modification of Diet in Renal Disease (MDRD) formula ^48^. *KDOQI*: the Kidney Disease Outcomes Quality Initiative (KDOQI) of chronic kidney disease (CKD) classification based on estimated glomerular filtration rate using the MDRD formula^49^ ; BMI: body mass index in kg/m^3^ ; *BMI* (*WHO*): BMI categories according to the World Health Organization^50^; *Diabetes*: diabetes status based on questionnaire, diagnosis by a medical doctor, or based on diabetic medication; *Hypertension*: hypertension status based on questionnaire, diagnosis by a medical doctor, or based on hypertensive medication (*antihypertensives*); *Antiplatelets*: usage of aspirin, dipyridamole and/or ADP inhibitor; *Lipid-lowering drugs* include statins, fibrates and other lipid-lowering medication; *Symptoms* prior to surgery; *Restenosis* indicates whether the CEA was de novo, or after a restenosis.

SMR analyses in the plaque were hindered by weak SNP-to-gene associations, leading to inclusion of only 15 genes (see **Figure 5**), with the majority not having enough SNPs available to conduct HEIDI. For these 15 genes, no significant SMR estimates were found at FDR < 0.1 (see **Supplemental Table 11**). **Figure 5** shows one nominally significant estimate (beta=0.056, SE=0.024, p=0.013) for the gene *ZFAND2A*, which passes the HEIDI test (15 SNPs, p=0.87), indicating that the association is unlikely to be explained by linkage.

**Figure 5.**
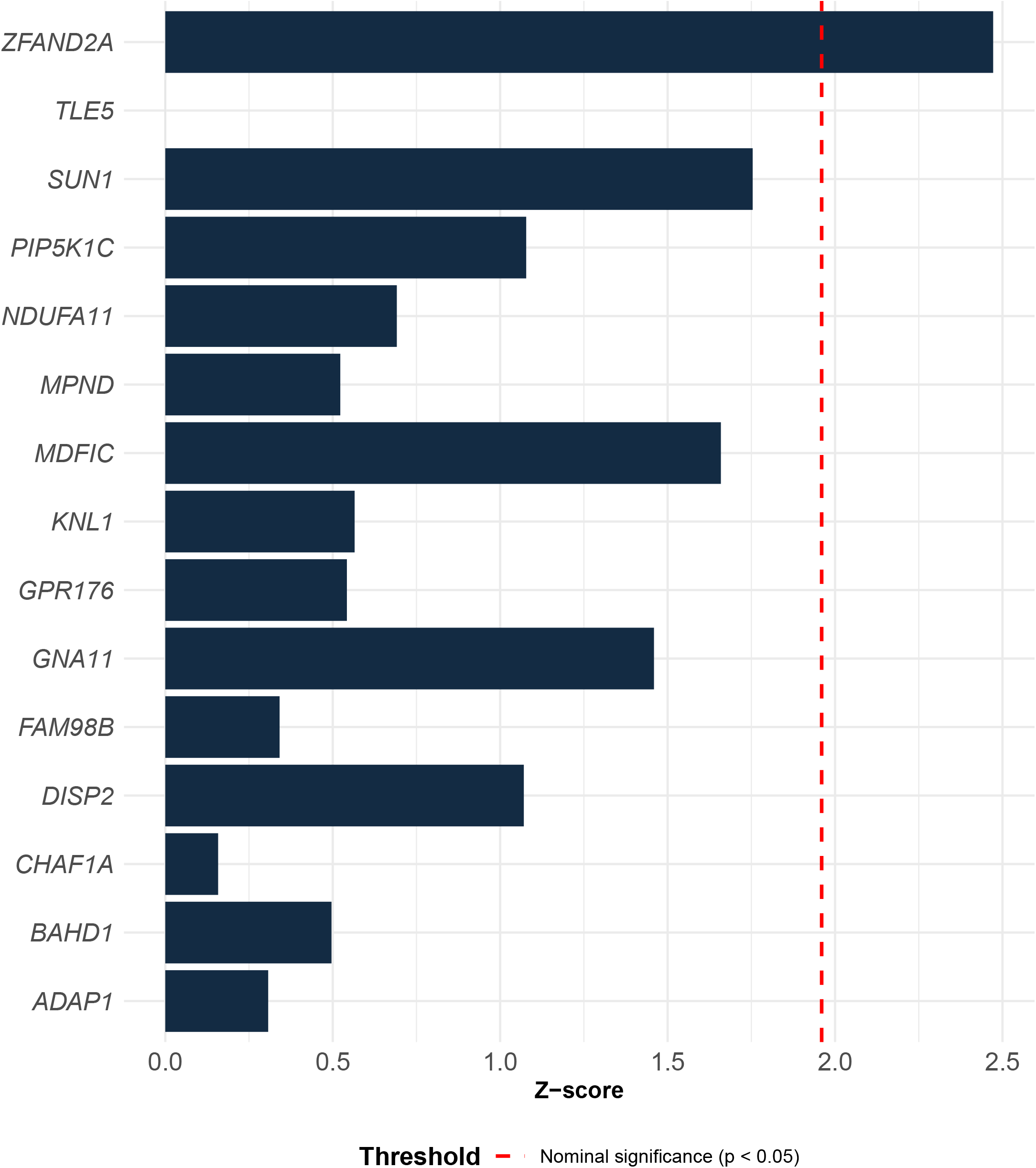
Causal estimates of gene expression in atherosclerotic plaques affecting MD. SMR results for all genes included in the analysis of atherosclerotic plaques. *Z-score*: SMR beta divided by standard error; no adjustment for multiple testing. For full results refer to Supplemental Table 11.

Given weak eQTL for the atherosclerotic plaque, we conducted SMR analyses of the effect of gene expression on CIMT, stroke and CAD to provide a positive control. This analysis yielded no nominally significant estimates (see **Supplemental Tables 12-14**), indicating that plaque eQTL data may not reflect genetic propensity for atherosclerosis, thus reducing the chance of finding significant effects on MD.

## Discussion

We examined which genetic risk factors are shared by major depression and atherosclerotic disease, and whether these genetic risk factors link to altered gene expression in various tissues, including atherosclerotic plaques. We found a causal connection between genetic liability for major depression and (ischemic) stroke, small vessel disease, and coronary artery disease, traceable to eight specific genomic regions. Altered expression of shared genetic factors was found to link to major depression predominantly in whole blood, brain, and heart, with limited evidence in atherosclerotic plaques.

First, using results from the latest GWAS, we showed that increased MD genetic liability was linked to a higher risk of developing (ischemic) stroke, small vessel disease, and CAD, but not vice versa. This causal direction, from depression to atherosclerotic disease, is in line with previous MR research^8,9^. Earlier studies in smaller GWAS (<500,000 for atherosclerotic diseases), found significant effects of depression genetic liability on CAD and small vessel disease, a pattern that our results from larger GWAS extend to (ischemic) stroke in general.

MR estimates suggest causal effects only in the absence of horizontal pleiotropy – SNPs having independent effects on two traits rather than affecting the outcome via the exposure. Here, follow-up analyses did not show evidence for horizontal pleiotropy for stroke – in contrast to CAD. This can likely be explained by the fact that CAD is a particularly broad and polygenic trait connected to multiple causal pathways. An effect of depression on CAD remains plausible in the context of results from other study designs, for example increased risk of CAD following depression has also been indicated by numerous prospective cohort studies^29^.

Next, we investigated causal relationships on the level of genomic regions. A high probability for containing a shared causal variant for MD and (ischemic) stroke or CAD was found for eight regions, with many of them containing protein-coding genes with known functions potentially relevant to etiology and pathogenesis of depression and/or atherosclerosis. For example, the gene *TMEM106B*, located in a region on chromosome that stood out for linking MD with multiple atherosclerotic traits, has been linked to central nervous system inflammation^30^. Accordingly, its association with neurodegenerative diseases may arise via regulation of microglia^31^. On the cellular level, *TMEM106B* is critical for the functioning of lysosomes^32^, a key player in plaque formation^33^. Whether lysosome activity may also be linked to depression is an open question, and the possibility of pleiotropy in the pathways remains.

Besides providing some fine-mapping information, colocalization was used to prioritize shared genes for depression and atherosclerotic disease that were promising candidates for downstream gene expression analyses. Based on this, we examined whether shared genetic vulnerability was related to gene expression in various tissues potentially influencing MD etiology. Expression – in blood, brain, heart, and assorted other tissues – of 28 genes from the shared causal regions with atherosclerosis, was indeed found to be potentially causal for depression. SMR analysis identified more than 20 links of gene expression with MD, spread across all investigated tissues (blood, brain, heart, artery, adipose). Some of these genes are known to have functions related to plausible shared mechanisms of MD and atherosclerotic diseases, such as immunometabolic processes like inflammation, lipid metabolism, and mitochondrial function:

For instance, *ZFAND2A* – with expression in atherosclerotic plaque, as well as whole blood, adipose tissue, atrial appendage, and left ventricle implicated here – is a zinc finger gene, a family of genes that has been found associated with metabolic dysregulation, in particular with glucose metabolism^34^. Whilst we might speculate about a link to inflammatory traits such as immunometabolic depression and atherosclerosis, further research would be required to elucidate the role of *ZFAND2A* in human health. A gene more directly linked to immune processes is *RASGRP2*, whose expression in brain tissue we found to be potentially causal for depression. *RASGRP2* is highly expressed in immune cells^35^ – specifically T cells^36^ – and linked to diabetes^37^. Relatedly, *GNA12*, here relevant to depression when expressed in whole blood or heart tissue, appears to be involved in regulation of pro-inflammatory cytokines IL-6 and IL-8^38^.

Lipid metabolism in particular is relevant to both depression and atherosclerotic diseases. In relation to this, *PLIN3* has been investigated for its role in macrophage lipid accumulation^33^, a contributor to atherosclerosis. Generally, *PLIN3* is involved in formation and function of lipid droplets^33^, organelles found abundantly in adipose tissues. Thus, the effect of *PLIN3* expression in visceral adipose tissues on depression found here may hint that the precise processes of lipid storage in adipose tissue should be considered relevant to the comorbidity.

Alterations in *GPR146* expression, coding for G-protein coupled receptor 146, are linked to cholesterol levels. Specifically, deficiency of *GPR146* leads to reduced LDL cholesterol and protection against hypercholesterolemia and atherosclerosis^39^. However, the current study shows an effect of *GPR146* in the cerebellum, a connection that has not been investigated before.

The gene *MRM2*, which in the present results stands out for significant causal estimates of its expression in most tissues, is connected to mitochondrial function^40^ and specifically altered in atherosclerosis^41^. Mitochondrial dysfunction seems to play a role in psychiatric disorders^41,42^, although details remain to be clarified.

To summarize, specific clues about the shared biology of depression and atherosclerosis can be derived from our results. Given the indirect nature of our genomics-based search, these functional descriptions must be interpreted as preliminary directions for future research rather than definite mechanistic explanations.

Finally, we were specifically interested in whether the shared genetic vulnerability acts through gene expression in the atherosclerotic plaque. However, we did not find strong evidence for alteration in gene expression in atherosclerotic plaque as a potential causal mechanism in depression.

In the analysis of atherosclerotic plaque eQTL we found little evidence for a causal effect on MD, with nominally significant (p < 0.05) estimates for expression of only one gene, *ZFAND2A*, and no significant estimates after adjusting for multiple testing. While evidence for involvement of *ZFAND2A* converges with our results from other tissues, in particular whole blood and heart, replication of this result is needed to reduce the likelihood of a false positive.

To further characterize the plaque eQTL, we tested their effect on traits known to be based on the occurrence of atherosclerotic plaque: CIMT, stroke, CAD. The results of this analysis were negative, indicating no causal effect of plaque eQTL on atherosclerotic disease. However, eQTL analysis provides an incomplete picture of the role of plaques in disease and comorbidity. GWAS-eQTL links are generally less abundant than expected^43^, and heterogeneity between the GWAS and plaque eQTL samples, in terms of age, health, and other covariates, may reduce a possible association. Further insights could be gained by analyzing additional omics layers, for instance proteomics.

Alternatively, the sparse findings may be due to the analysis being underpowered for the plaque samples. Whilst a comparison to the sample sizes for tissues with significant causal estimates does not suggest a substantial contrast, power may nevertheless be lower if effect sizes in the eQTL are smaller in plaque tissue. Similarly, standard errors in plaque eQTL can be relatively large, given that despite the homogenous cohort – patients with clinical symptoms at an advanced stage of disease – plaque samples are heterogenous in nature.

The current study benefits from several factors. In selecting GWAS we relied on the largest available meta-analyses for all traits, yielding the most robust results. Additionally, we were, for the first time, able to investigate eQTL in the culprit tissue for atherosclerotic diseases with regard to depression. MR and colocalization complement each other – whilst relying on distinct methodologies, both examine the shared causal basis of two traits – thereby providing a layer of triangulation that enhances robustness of the study^44^.

Nevertheless, the current study is subject to several limitations. All selected GWAS meta-analysis samples were limited to European ancestry, reducing generalizability to other ancestries. Additionally, heterogeneity in the GWAS phenotypes may have diluted relevant signals, especially for MD. The MD GWAS meta-analysis is based on a broad phenotype ranging from clinical diagnosis to simple self-report questions^10^, likely capturing less specific genetic architecture^45^. Relatedly, the clinical heterogeneity of major depressive disorder, also reflected at the genetic level^46^, may obscure the applicability of findings that are potentially driven by a biologically distinct subgroup of patients exhibiting immuno-metabolic alterations.

## Conclusion

This study indicates that part of the connection between depression and atherosclerotic disease stems from a shared genetic root. The genetic liability for major depression encompasses mechanisms likely causal for the development of coronary artery disease and stroke.

The study pinpointed eight regions in the genome harboring potentially causal variants shared by depression with coronary artery disease and stroke. In these shared genomic regions, pathways acting through the expression of 28 genes involved in immuno-metabolic pathways in blood, brain and heart are likely involved in the pathophysiology of depression. Less consistent evidence was found for similar mechanisms acting in atherosclerotic plaques.

Overall, the present study identified specific shared biological pathways potentially underlying the established risk of atherosclerotic disease in depression. For clinical practice, evidence of shared biological mechanisms suggests the relevance of carefully monitoring cardiometabolic health and atherosclerotic risk in subjects with depression. Furthermore, pathways identified in the present study could guide the development of new treatments to prevent depression and its heightened atherosclerotic risk.

## Data Availability

The data, analytic methods, and study materials are available to other researchers. The Athero-Express Biobank Study data are available through DataverseNL; the other datasets are available through the respective sources as reported in Supplemental Table 1 and 2. The main scripts used for the quality control and the (meta-)analysis of the data are available through GitHub.

https://doi.org/10.34894/4IKE3T

https://github.com/CirculatoryHealth/MR_CVD_MDD

## Acknowledgements

We are thankful for the support of the Leducq Foundation ‘PlaqOmics’ and ‘AtheroGen’, and the Chan Zuckerberg Initiative ‘MetaPlaq’. The collaborative project ‘Getting the Perfect Image’ was co-financed through use of PPP Allowance awarded by Health∼Holland, Top Sector Life Sciences & Health, to stimulate public-private partnerships.

Claudia Tersteeg, Krista den Ouden, Mirjam B. Smeets, and Loes B. Collé are graciously acknowledged for their work on the DNA extraction. Astrid E.M.W. Willems, Evelyn Velema, Kristy M. J. Vons, Sara Bregman, Timo R. ten Brinke, Sara van Laar, Louise M. Catanzariti, Joyce E.P. Vrijenhoek, Sander M. van de Weg, Arjan H. Schoneveld, Arnold Koekman, Arjan Boltjes, Petra H. Homoed-van der Kraak, and Aryan Vink are graciously acknowledged for their past and continuing work on the Athero-Express Biobank Study. We would also like to thank all the (former) employees involved in the Athero-Express Biobank Study of the Departments of Surgery of the St. Antonius Hospital Nieuwegein and University Medical Center Utrecht for their continuing work. Lastly, we would like to thank all participants of the Athero-Express Biobank Study; without you these kinds of studies would not be possible.

We would also like to thank research participants and employees of 23andMe, Inc. for making this work possible.

## Disclosures

### Data and code availability

The data, analytic methods, and study materials are available to other researchers. The Athero-Express Biobank Study data are available through DataverseNL (https://doi.org/10.34894/4IKE3T); the other datasets are available through the respective sources as reported in **Supplemental Table 1** and **2**. The main scripts used for the quality control and the (meta-)analysis of the data are available through GitHub (https://github.com/CirculatoryHealth/MR_CVD_MDD).

