## Supplemental Materials for "Major depression and atherosclerotic disease: Linking shared genetics to pathways in blood, brain, heart, and atherosclerotic plaques"

### Supplemental Methods

#### GWAS summary statistics

Summary statistics were preprocessed by selecting only single-nucleotide polymorphisms (SNPs) present in the 1000 Genomes European ancestry reference (phase 3, version 5) and, where absent, assigning rsIDs based on genome build b37^1^.

#### Genetic correlations

The high-definition likelihood method (HDL) extends the established linkage disequilibrium score regression method (LDSC) by considering LD across nearly the entire autosomal genome, resulting in a lower variance of estimates and greater power to identify significant genetic correlations. Here, we calculated genetic correlations between MD and eight different atherosclerotic phenotypes using the pre-computed UK Biobank reference panel with 1,029,876 imputed HapMap3 SNPs as prepared for the HDL method ([https://github.com/zhenin/HDL/](https://github.com/zhenin/HDL/)2))^2^. As recommended for linkage disequilibrium score regression (LDSC), traits with a z-score of the heritability < 4 were excluded from this analysis^3^.

#### Mendelian Randomization

The main Mendelian Randomization analysis was repeated once for a potential causal effect of depression on atherosclerosis (exposure: MD, outcomes: ALLSTROKE, IS, SVD, LAS, CES, CAC, CIMT), and once for a potential causal effect of atherosclerosis on depression (exposures: ALLSTROKE, IS, SVD, LAS, CES, CAC, CIMT, outcome: MD).

For each phenotype combination, instruments were selected by filtering the exposure data by minor allele frequency (maf) > 0.01 and p-value < 5x10^-8^ and then clumping them with a kb window of 10,000 and R² = 0.001 (default) using the 1000 Genomes European ancestry LD reference (phase 3, version 5)^1^. We assessed the strength of the instruments’ association with the exposure by calculating F-statistics for all SNPs, with F > 10 indicating adequate instrument strength^4^.

Here, we focus our attention on those phenotypes with significant causal estimates, which are the ones relevant to further analyses. For these phenotypes, the robustness of these estimates was tested by employing related methods that allow for violations to the central assumptions of MR: Weighted median causal estimates are robust to up to half the SNPs being invalid instruments^5^. MR Egger causal estimates are robust to all SNPs being invalid instruments under the so-called INSIDE assumption - the strength of their association with the exposure must be independent of their direct effect on the outcome^6^.

Heterogeneity in the SNP effects, which may indicate presence of pleiotropy, was examined using Cochran’s Q test, as well as leave-one-out and single SNP plots. The presence of horizontal pleiotropy, SNPs having independent effects on both phenotypes, was investigated using the MR Egger intercept and the MR pleiotropy residual sum and outlier (MR-PRESSO) test. As the MR Egger intercept may be interpreted as the average of the pleiotropic effects across the genetic instruments, an estimate that differs significantly from 0 indicates pleiotropy^6^.

If a global test for horizontal pleiotropy, based on the residual sum of squares, is positive, MR-PRESSO detects and removes horizontal pleiotropic outliers and assesses whether this correction significantly distorts causal estimates^7^.

#### Athero-Express Biobank Study

The Athero-Express Biobank Study (AE) is an ongoing longitudinal biobank study including patients that undergo arterial endarterectomy in two Dutch tertiary referral centers since 2002^8^. For the present study, subsequent patients were included who underwent an endarterectomy of either the carotid or the femoral and iliac arteries and of which genotyping and transcriptomic data were available. Clinical data were extracted from patient medical files and standardized questionnaires. This study complies with the Declaration of Helsinki, and all participants provided informed consent. The medical ethical committees of the respective hospitals approved this study registered under number 22/018.

##### DNA isolation, genotyping, and imputation

**DNA isolation and genotyping**

As described in the study design of the AE blood samples were obtained prior to surgery and stored at -80℃^8^. Carotid plaque specimens were removed during surgery and immediately processed in the laboratory. Specimens were cut transversely into segments of 5 mm. The culprit lesion (the region with most severe stenosis) was identified, fixed in 4% formaldehyde, embedded in paraffin, and processed for histological examination^8^. Remaining segments were stored at -80℃.

We genotyped the AE in three separate, but consecutive experiments^9^. In short, DNA was extracted from EDTA blood or (when no blood was available) plaque samples of 1,858 consecutive patients from the Athero-Express Biobank Study and genotyped in 3 batches. For the Athero-Express Genomics Study 1 (AEGS1) 891 patients (602 males, 262 females, 27 unknown sex), included between 2002 and 2007, were genotyped (440,763 markers) using the Affymetrix Genome-Wide Human SNP Array 5.0 (SNP5) chip (Affymetrix Inc., Santa Clara, CA, USA) at Eurofins Genomics(<https://www.eurofinsgenomics.eu/>, formerly known as AROS). For the Athero-Express Genomics Study 2 (AEGS2) 954 patients (640 mal~~k~~es, 313 females, 1 unknown sex), included between 2002 and 2013, were genotyped (587,351 markers) using the Affymetrix AxiomⓇ GW CEU 1 Array (AxM) at the Genome Analysis Center ([https://www.helmholtz-muenchen.de](https://www.helmholtz-muenchen.de/)). The two first batches, AEGS1 and AEGS2, were described before^9^. For the Athero-Express Genomics Study 3 (AEGS3) 658 patients (448 males, 203 females, 5 unknown sex), included between 2002 and 2016, were genotyped (693,931 markers) using the Illumina GSA MD v1 BeadArray (GSA) at Human Genomics Facility, HUGE-F (http://glimdna.org/index.html). All experiments were carried out according to OECD standards. We used the genotyping calling algorithms as advised by Affymetrix (AEGS1 and AEGS2) and Illumina (AEGS3): BRLMM-P, AxiomGT1, and Illumina GenomeStudio respectively.

**Quality control after genotyping**

After genotype calling, we adhered to community standard quality control and assurance (QCA) procedures of the genotype data from AEGS1, AEGS2, and AEGS3^9^. Samples with low average genotype calling and sex discrepancies (compared to the clinical data available) were excluded. The data was further filtered on 1) individual (sample) call rate > 97%, 2) SNP call rate > 97%, 3) minor allele frequencies (MAF) > 3%, 4) average heterozygosity rate ± 3.0 standard deviation 5) relatedness (pi-hat > 0.20), 6) Hardy–Weinberg Equilibrium (HWE p < 1.0×10^−3^), and 7) Monomorphic SNPs (< 1.0×10^−6^). Principal component analysis (PCA) was applied to determine genetic ancestry using 26 populations from the 1000G phase 3 (version 5) as a reference^33^. After QCA 2,493 samples remained, 108 of non-European descent/ancestry, and 156 related pairs. These comprise 890 samples and 407,712 SNPs in AEGS1, 869 samples and 534,508 SNPs in AEGS2, and 649 samples and 534,508 SNPs in AEGS3 remained.

**Imputation**

Before phasing using SHAPEIT2, data was lifted to genome build b37 using the liftOver tool from UCSC (<https://genome.ucsc.edu/cgi-bin/hgLiftOver>). Finally, data was imputed with 1000G phase 3, version 5 and HRC release 1.1 as a reference using the Michigan Imputation Server (<https://imputationserver.sph.umich.edu/>)^10^. These results were further integrated using QCTOOL v2, where HRC imputed variants are given precedence over 1000G phase 3 imputed variants.

**RNA isolation and RNA sequencing**

**RNA isolation and library preparation**

A total of 700 segments were selected from patients who were included in the study between 2002 and 2016^11^. As the RNA isolated from the archived advanced atherosclerotic lesion is fragmented, we have ultimately employed the CEL-seq2 method^12^. CEL-seq2 yielded the highest mappability reads to the annotated genes compared to other library preparation protocols. The methodology captures 3’-end of polyadenylated RNA species and includes unique molecular identifiers (UMIs), which allow direct counting of unique RNA molecules in each sample.

**Sequencing read mapping and quality filtering**

Libraries were sequenced on the Illumina Nextseq500 platform; a high output paired-end run of 2 × 75 bp was performed (Utrecht Sequencing Facility). The reads were demultiplexed and aligned to human cDNA reference (Ensembl 84) using the BWA (0.7.13). Multiple reads mapping to the same gene with the same unique molecular identifier (UMI, 6bp long) were counted as a single read. The raw read counts were corrected for UMI sampling (corrected_count=-4096*(ln(1-(raw_count/4096)))), normalized for sequencing depth and quantile normalized (core scripts can be found in <https://github.com/mmokry/bulkCEL-seq2> and <https://github.com/mmokry/seurat_meets_bulk_AE>). We have detected a median of 19.501 (SD = 5.874) genes per sample with at least one unique read, and discarded samples (n=46) with less than 9,000 detected genes from further analysis. For all the subsequent analyses, we have excluded all the ribosomal genes and used only the protein-coding genes with annotated HGCN names.

**Expression quantitative trait loci analysis**

First, the UMI corrected RNAseq counts were mapped to corresponding hg19 biomart gene information; non-protein coding genes were excluded as well as those lying on non-standard (alternative) chromosomes. UMI-corrected counts reported as infinite float values were replaced by the largest observed finite count value. Next, a sweep over a missingness threshold from 10% to 100% in steps of 10% was conducted, and a separate gene dataset was prepared for each threshold. The filter is applied by removing genes with zero counts for more than the threshold-portion of samples. TMM normalization was applied as provided by the conorm package and inverse normal transform normalization using the scipy.stats package in Python (version 3.4.3)^13^.

For these eQTL analyses we followed (parts of) the GTEx V7 methods (<https://storage.googleapis.com/gtex-public-data/Portal_Analysis_Methods_v7_09052017.pdf>). To determine the number of expression principal components (PCs) to correct for subtle substructure in the expression data, the following process was employed. For sample covariates and subsequent sample exclusion, 2 genetics principal components (PCs) and 100 expression PCs were calculated. For expression data, randomized truncated principal component analysis (PCA) estimation was used due to the large dataset^14^. For the first 2 expression covariates, the sample Mahalanobis distance was calculated, and samples below the Chi^2^ (n=3, alpha=0.95) threshold were selected, similarly as described for GTEx V7. Then a sweep over the amount of included expression PCs was performed from n=0 to 100 components and each total (2+n) covariates was saved to a different file. QTLtools^15^ was used to select the missingness threshold (10-100%) of the gene counts and expression covariate counts (0-100) from all generated combinations for subsequent analysis. For this, SNPs were filtered on MAF larger than 0.03 and INFO score larger than 0.4 and stored as VCF-file as required for 80% power. QTLtools was run in cis permutation mode (100-10,000x) with a window of 1Mb and the amount of gene-level and genome-wide level results from the permutation test adjusted p-values was determined. Here it was found that the amount of genome-wide significant results flattens at a missingness value of 50%, where then a peak is found for 45 expression covariates.

For the final *cis*-acting eQTL mapping we employed the TensorQTL^16^ package on an Nvidia RTX6000 GPU. Based on the above, we included 2 genetic principal components (PCs) and 45 expression PCs for the eQTL analyses and used a missingness 50% threshold for the gene counts. We found that these 2 genetic PCs and 45 expression PCs also capture age and sex related information, no correction for these confounders was applied. To boost power, we did not exclude samples based on the genetic PCA, rather chose to correct for this by adding the first 2 PCs (which explain most of the variation with respect to the genetic substructure). For the *cis*-eQTL mapping, the above generated VCF-files (MAF > 0.03, INFO > 0.4) were converted to PLINK^17^ BED-file format using the default settings of PLINK2 (2.0.0a2lm, <https://www.cog-genomics.org/plink/2.0/>) and as required by TensorQTL. The total sample size of included patients was 626 (AEGS1 n=268, AEGS n=345, AEGS3 n=13), with 614 carotid endarterectomy (98%) and 12 other (external carotid and femoral) endarterectomy patients, and 11 (1.6%) were from non-European genetic ancestry based on PCA.

### Supplemental Tables

###### **Supplemental** **Table 1:** Overview of GWAS Summary Statistics Included in this Study.

###### Largest GWAS meta-analyses to date were identified for each trait. Cases: number of cases for disease phenotypes; Total Sample Size: Total number of all included GWAS’ European ancestry participants; n.a.: not applicable.

| Phenotype | Abbreviation | Cases | Total Sample Size | Consortium | Reference |
| --- | --- | --- | --- | --- | --- |
| Major Depression | MD/DEP | 525,197 | 3,887,532 | PGC | Adams et al., 2025^18^ |
| Any stroke | ALLSTROKE | 73,652 | 1,308,460 | GIGASTROKE | Mishra et al., 2022^19^ |
| Any Ischemic stroke | IS | 62,100 | 1,296,908 | GIGASTROKE | Mishra et al., 2022^19^ |
| Cardio- Embolic stroke | CES | 10,804 | 1,245,612 | GIGASTROKE | Mishra et al., 2022^19^ |
| Small Vessel Disease | SVD | 6811 | 1,241,619 | GIGASTROKE | Mishra et al., 2022^19^ |
| Large Artery Stroke | LAS | 6399 | 1,241,207 | GIGASTROKE | Mishra et al., 2022^19^ |
| Coronary Artery Disease | CAD | 117,010 | 1,165,690 | Millionhearts | Aragam et. al., 2022^20^ |
| Carotid Intima-Media Thickness | CIMT | n.a. | 71,128 |  | Franceschini et al. 2018^21^ |
| Coronary Artery Calcification | CAC | n.a. | 26,909 |  | Kavousi et al., 2023^22^ |

###### **Supplemental Table 2:** References and sample sizes of eQTL datasets used for SMR analysis of gene expression in blood/brain/heart.

Datasets selected from the SMR-Portal database (<https://yanglab.westlake.edu.cn/smr-portal/doc/about#about_smr_database>).

| QTL Study | Label | Sample size | Tissue A | Tissue B | Reference | URL |
| --- | --- | --- | --- | --- | --- | --- |
| eQTLGen | eQTLGen | 31,684 | Blood | Blood | Võsa, U. et al. Nat Genet. 2021^23^ | https://eqtlgen.org/ |
| BrainMeta | eQTL_BrainMeta | 2,865 | Brain | Brain | Qi, T. et al. Nat Genet. 2022^24^ | <https://yanglab.westlake.edu.cn/data/brainmeta/cis_eqtl/> |
| GTEx v8 eQTL | eQTL_GTEx_Adipose_Visceral_Omentum | 541 | Adipose | Adipose Visceral Omentum | GTEx Consortium. Science. 2020^25^ | https://www.gtexportal.org/home/ |
|  | eQTL_GTEx_Artery_Aorta | 432 | Blood Vessel | Artery Aorta |  |  |
|  | eQTL_GTEx_Artery_Coronary | 240 | Blood Vessel | Artery Coronary |  |  |
|  | eQTL_GTEx_Brain_Amygdala | 152 | Brain | Brain Amygdala |  |  |
|  | eQTL_GTEx_Brain_Anterior_cingulate_cortex_BA24 | 176 | Brain | Brain Anterior cingulate cortex BA24 |  |  |
|  | eQTL_GTEx_Brain_Caudate_basal_ganglia | 246 | Brain | Brain Caudate basal ganglia |  |  |
|  | eQTL_GTEx_Brain_Cerebellar_Hemisphere | 215 | Brain | Brain Cerebellar Hemisphere |  |  |
|  | eQTL_GTEx_Brain_Cerebellum | 241 | Brain | Brain Cerebellum |  |  |
|  | eQTL_GTEx_Brain_Cortex | 255 | Brain | Brain Cortex |  |  |
|  | eQTL_GTEx_Brain_Frontal_Cortex_BA9 | 209 | Brain | Brain Frontal Cortex BA9 |  |  |
|  | eQTL_GTEx_Brain_Hippocampus | 197 | Brain | Brain Hippocampus |  |  |
|  | eQTL_GTEx_Brain_Hypothalamus | 202 | Brain | Brain Hypothalamus |  |  |
|  | eQTL_GTEx_Brain_Nucleus_accumbens_basal_ganglia | 246 | Brain | Brain Nucleus accumbens basal ganglia |  |  |
|  | eQTL_GTEx_Brain_Putamen_basal_ganglia | 205 | Brain | Brain Putamen basal ganglia |  |  |
|  | eQTL_GTEx_Brain_Spinal_cord_cervical_c-1 | 159 | Brain | Brain Spinal cord cervical c-1 |  |  |
|  | eQTL_GTEx_Brain_Substantia_nigra | 139 | Brain | Brain Substantia nigra |  |  |
|  | eQTL_GTEx_Heart_Atrial_Appendage | 429 | Heart | Heart Atrial Appendage |  |  |
|  | eQTL_GTEx_Heart_Left_Ventricle | 432 | Heart | Heart Left Ventricle |  |  |

###### **Supplemental Table 3:** Genetic correlations between depression and atherosclerotic phenotypes.

Genetic correlations between depression and atherosclerotic phenotypes are displayed (HDL estimates with standard errors in brackets). LAS was not considered, due to a heritability z-score < 4 (heritability estimate divided by standard error), indicating insufficient statistical power to calculate genetic correlations.

*ALLSTROKE*: any stroke; *IS*: ischemic stroke; *CES*: cardioembolic stroke; *LAS*: large artery stroke; *SVD*: small vessel disease; *CAD*: coronary artery disease; *CAC*: coronary artery calcification; *CIMT*: carotid intima-media thickness; *DEP*: major depression; *n.a.*: not applicable; * Indicates significance at p < 0.05/8.

|  | **Heritability z-score** | **Genetic correlation with DEP** | **SE** | **P-value** |
| --- | --- | --- | --- | --- |
| **ALLSTROKE** | 11.2 | 0.193 | 0.027 | 1.2x10^12^* |
| **IS** | 11.89 | 0.203 | 0.028 | 4.71x10^13^* |
| **CES** | 5.95 | 0.055 | 0.035 | 1.15x10^1^ |
| **LAS** | 2.51 | 0.158 | 0.063 | 1.19x10^2^ |
| **SVD** | 4.8 | 0.242 | 0.054 | 7.4x10^6^* |
| **CAD** | 14.37 | 0.232 | 0.02 | 6.0x10^32^* |
| **CAC** | 5.71 | 0.081 | 0.03 | 6.87x10^3^ |
| **CIMT** | 6.2 | 0.035 | 0.039 | 3.69x10^1^ |

**Supplemental Table 4:** MR of the effect of genetically predicted depression on atherosclerotic phenotypes.

Main MR results from inverse-variance weighted (IVW) method, with weighted mean (WM) and MR Egger as additional sensitivity analyses. *N SNPs*: number of SNPs included as instrumental variable; *OR:* Odds ratio per doubling (2-fold increase) in the prevalence of the exposure, obtained by multiplying the causal estimate/beta by ln(2) as suggested by^26^; CI: 95% confidence interval; *ALLSTROKE*: any stroke; *IS*: ischemic stroke; *CES*: cardioembolic stroke; *LAS*: large artery stroke; *SVD*: small vessel disease; *CAD*: coronary artery disease; *CAC*: coronary artery calcification; *CIMT*: carotid intima-media thickness; *DEP*: major depression; *** Indicates p-value significant at False Discovery Rate < 0.1.

|  |  |  | IVW | | WM | | MR Egger | |
| --- | --- | --- | --- | --- | --- | --- | --- | --- |
| Outcome | **Exposure** | **N SNPs** | **OR (CI)** | **P-value** | **OR (CI)** | **P-value** | **OR (CI)** | **P-value** |
| ALLSTROKE | DEP | 362 | 1.1 (1.05-1.16) | 9.47x10^8^* | 1.09 (1.03-1.16) | 7.39x10^5^* | 1.22 (0.98-1.51) | 0.011 |
| IS | DEP | 362 | 1.11 (1.05-1.17) | 1.52x10^7^* | 1.09 (1.02-1.17) | 2.97x10^4^* | 1.16 (0.91-1.46) | 0.082 |
| CES | DEP | 360 | 1.07 (0.96-1.19) | 0.069 | 1.04 (0.9-1.22) | 4.30x10^1^ | 1.24 (0.77-1.98) | 0.206 |
| LAS | DEP | 358 | 1.09 (0.95-1.26) | 0.073 | 1.09 (0.89-1.33) | 2.48x10^1^ | 1.8 (0.95-3.41) | 0.01* |
| SVD | DEP | 358 | 1.23 (1.06-1.41) | 4.76x10^5^* | 1.3 (1.07-1.57) | 1.18x10^4^* | 1.59 (0.85-2.98) | 0.035* |
| CAD | DEP | 348 | 1.2 (1.13-1.26) | 3.76x10^22^* | 1.19 (1.13-1.26) | 7.90x10^23^* | 1.34 (1.07-1.68) | 2.43x10^4^* |
| CAC | DEP | 365 | 1.04 (0.92-1.18) | 0.338 | 1.01 (0.85-1.19) | 0.9 | 0.99 (0.58-1.72) | 0.978 |
| CIMT | DEP | 365 | 1 (1-1.01) | 0.828 | 1 (0.99-1.01) | 1.00 | 1 (0.98-1.02) | 0.781 |

###### **Supplemental Table 5:** Instrument strength in MR with exposure depression.

Range of F-statistics of the association between SNPs and exposure. F-statistic approximated as est^2^/se^2^ according to Pierce *et al*^4^. F > 10 indicates sufficient strength of the instrumental variable. *N SNPs*: number of SNPs included as instrumental variable; *ALLSTROKE*: any stroke; *IS*: ischemic stroke; *CES*: cardioembolic strokes; *LAS*: large artery stroke; *SVD*: small vessel disease; *CAD*: coronary artery disease; *CAC*: coronary artery calcification; *CIMT*: carotid intima-media thickness. Note that SVD was not included as an exposure, as no valid instruments were identified (N SNPs = 0).

| Outcome | Exposure | N SNPs | Min. F-statistics | Max. F-statistics |
| --- | --- | --- | --- | --- |
| ALLSTROKE | DEP | 362 | 28.9 | 198.27 |
| IS | DEP | 362 | 28.9 | 198.27 |
| CES | DEP | 360 | 28.9 | 198.27 |
| LAS | DEP | 358 | 28.9 | 198.27 |
| SVD | DEP | 358 | 28.9 | 198.27 |
| CAD | DEP | 348 | 28.9 | 197.11 |
| CAC | DEP | 365 | 28.9 | 198.27 |
| CIMT | DEP | 365 | 28.9 | 198.27 |
| DEP | ALLSTROKE | 23 | 29.6 | 84.0 |
| DEP | IS | 23 | 30.0 | 83.6 |
| DEP | CES | 7 | 31.5 | 212.7 |
| DEP | LAS | 3 | 30.5 | 41.2 |
| DEP | SVD | 173 | 29.9 | 1065.0 |
| DEP | CAD | 6 | 34.0 | 202.2 |
| DEP | CAC | 8 | 27.6 | 62.9 |

###### **Supplemental Table 6:** Cochran’s Q for instrument heterogeneity

Cochran’s Q estimator for inverse variance weighted method and MR Egger method. A significant Cochran’s Q indicates the presence of an outlier. *df*: degrees of freedom. *ALLSTROKE*: any stroke; *IS*: ischemic stroke; *CES*: cardioembolic stroke; *LAS*: large artery stroke; *SVD*: small vessel disease; *CAD*: coronary artery disease; *CAC*: coronary artery calcification; *CIMT*: carotid intima-media thickness; *DEP*: major depression.

|  | | Cochran's Q | | | | | |
| --- | --- | --- | --- | --- | --- | --- | --- |
| Outcome | **Exposure** | **MR Egger** | | | **Inverse variance weighted** | | |
|  |  | **Q** | **df** | **P-value** | **Q** | **df** | **P-value** |
| ALLSTROKE | DEP | 567 | 360 | 1.75x10^11^ | 570 | 361 | 1.30x10^11^ |
| IS | DEP | 566 | 360 | 2.34x10^11^ | 566 | 361 | 2.71x10^11^ |
| CES | DEP | 423 | 358 | 0.010 | 424 | 359 | 0.010 |
| LAS | DEP | 408 | 356 | 0.029 | 414 | 357 | 0.020 |
| SVD | DEP | 470 | 356 | 4.75x10^5^ | 472 | 357 | 4.28x10^5^ |
| CAD | DEP | 983 | 346 | 1.94x10^62^ | 990 | 347 | 4.01x10^63^ |
| CAC | DEP | 393 | 363 | 0.134 | 393 | 364 | 0.142 |
| CIMT | DEP | 421 | 363 | 0.020 | 421 | 364 | 0.021 |

###### **Supplemental Table 7:** MR-Egger intercept to assess horizontal pleiotropy in forward MR.

A significant intercept of the MR-Egger method indicates presence of horizontal pleiotropy. *SE*: standard error; *ALLSTROKE*: any stroke; *IS*: ischemic stroke; *CES*: cardioembolic stroke; *LAS*: large artery stroke; *SVD*: small vessel disease; *CAD*: coronary artery disease; *CAC*: coronary artery calcification; *CIMT*: carotid intima-media thickness; *DEP*: major depression.

| Outcome | Exposure | Egger intercept | SE | P-value |
| --- | --- | --- | --- | --- |
| ALLSTROKE | DEP | -0.00282 | 0.00211 | 0.182 |
| IS | DEP | -0.00123 | 0.00229 | 0.593 |
| CES | DEP | -0.00400 | 0.00457 | 0.382 |
| LAS | DEP | -0.01389 | 0.00619 | 0.254 |
| SVD | DEP | -0.00738 | 0.00606 | 0.223 |
| CAD | DEP | -0.00329 | 0.00218 | 0.133 |
| CAC | DEP | 0.00133 | 0.00530 | 0.802 |
| CIMT | DEP | 0.00007 | 0.00020 | 0.736 |

###### **Supplemental Table 8:** MR PRESSO results for forward MR.

Results of global test evaluating the presence of horizontal pleiotropy, outlier-corrected test removing specific instrumental variables, and distortion test assessing whether original causal estimate is distorted by outlier variants. *RSS*: Residual sum of squares; *SE*: standard error; *ALLSTROKE*: any stroke; *IS*: ischemic stroke; *CES*: cardioembolic stroke; *LAS*: large artery stroke; *SVD*: small vessel disease; *CAD*: coronary artery disease; *CAC*: coronary artery calcification; *CIMT*: carotid intima-media thickness; *DEP*: major depression.

| Outcome | Exposure | N SNPs | Global Test | | Outlier Corrected | | | Distortion Test | |
| --- | --- | --- | --- | --- | --- | --- | --- | --- | --- |
|  |  |  | **RSS** | **P-value** | **Estimate** | **SE** | **P-value** | **Coefficient** | **P-value** |
| ALLSTROKE | DEP | 362 | 423.12 | 0.019 |  |  | NA |  |  |
| IS | DEP | 362 | 423.12 | 0.026 |  |  | NA |  |  |
| CES | DEP | 360 | 423.12 | 0.018 |  |  | NA |  |  |
| LAS | DEP | 358 | 423.12 | 0.027 | 0.00018 | 0.00237 | 0.938 | 181.2222 | 0.131 |
| SVD | DEP | 358 | 423.12 | 0.020 |  |  | NA |  |  |
| CAD | DEP | 348 | 423.12 | 0.019 | 0.00069 | 0.00235 | 0.768 | -25.0045 | 0.950 |
| CAC | DEP | 365 | 423.12 | 0.016 | 0.00018 | 0.00237 | 0.938 | 181.2222 | 0.119 |
| CIMT | DEP | 365 | 423.12 | 0.029 | 0.00078 | 0.00238 | 0.742 | -33.7153 | 0.947 |

###### **Supplemental Table 9:** MR of the effect of genetically predicted atherosclerotic phenotypes on depression.

Main MR results from inverse-variance weighted (IVW) method, with weighted mean (WM) and MR Egger as additional sensitivity analyses. *N SNPs*: number of SNPs included as instrumental variable; *OR:* Odds ratio per doubling (2-fold increase) in the prevalence of the exposure, obtained by multiplying the causal estimate/beta by ln(2) as suggested by^26^; CI: 95% confidence interval; *ALLSTROKE*: any stroke; *IS*: ischemic stroke; *CES*: cardioembolic stroke; *LAS*: large artery stroke; *SVD*: small vessel disease; *CAD*: coronary artery disease; *CAC*: coronary artery calcification; *CIMT*: carotid intima-media thickness; *DEP*: major depression; *** Indicates p-value significant at False Discovery Rate < 0.1.

|  |  |  | **IVW** | | **WM** | | **MR Egger** | |
| --- | --- | --- | --- | --- | --- | --- | --- | --- |
| **Outcome** | **Exposure** | **N SNPs** | **OR (CI)** | **P-value** | **OR (CI)** | **P-value** | **OR (CI)** | **P-value** |
| DEP | ALLSTROKE | 23 | 1.02 (0.98-1.06) | 0.193 | 1.01 (0.98-1.05) | 0.266 | 1.1 (0.89-1.34) | 0.216 |
| DEP | IS | 23 | 1.01 (0.97-1.06) | 0.418 | 1.01 (0.98-1.05) | 0.233 | 1.12 (0.91-1.37) | 0.137 |
| DEP | CES | 7 | 1.01 (0.98-1.03) | 0.496 | 1 (0.98-1.02) | 0.936 | 0.99 (0.93-1.05) | 0.714 |
| DEP | LAS | 3 | 1.01 (0.98-1.04) | 0.326 | 1.02 (0.99-1.05) | 0.114 | 0.94 (0.78-1.14) | 0.551 |
| DEP | CAD | 173 | 1 (0.99-1.02) | 0.631 | 1 (0.99-1.02) | 0.509 | 1 (0.96-1.03) | 0.736 |
| DEP | CAC | 6 | 1.01 (1-1.02) | 0.092 | 1.01 (0.99-1.02) | 0.178 | 0.99 (0.96-1.03) | 0.674 |
| DEP | CIMT | 8 | 0.81 (0.55-1.17) | 0.104 | 0.9 (0.55-1.46) | 0.531 | 0.98 (0.28-3.45) | 0.973 |

*Note*: As none of the main results were significant, no further sensitivity analyses are presented here.

###### **Supplemental Table 10:** Regions with high probability of colocalization with depression.

Top results of colocalization analysis by lead SNP. *Lead SNP*: SNP at the center of the tested region, selected for being significantly associated with MD; *Chr*: chromosome; *Bp*: base pair position; *N SNPs*: number of SNPs in region; Coloc hypotheses (here, trait 1 = major depression and trait 2 = atherosclerotic phenotype) were *H0*: no causal variant for either trait; *H1*: causal variant for trait 1; *H2*: posterior probability of causal variant for trait 2; *H3*: separate causal variants for both traits; *H4*: shared causal variant; *ALLSTROKE*: any stroke; *IS*: ischemic stroke; *CAD*: coronary artery disease.

|  |  |  |  |  | Posterior Probabilities | | | | |
| --- | --- | --- | --- | --- | --- | --- | --- | --- | --- |
| Phenotype | **Lead SNP** | **Chr** | **Bp** | **N SNPs** | **H0** | **H1** | **H2** | **H3** | **H4** |
| ALLSTROKE | rs10950392 | 7 | 12263538 | 7994 | 0.00 | 0.03 | 0.00 | 0.01 | 0.96 |
| ALLSTROKE | rs7351050 | 19 | 4044579 | 4262 | 0.00 | 0.07 | 0.00 | 0.04 | 0.88 |
| CAD | rs10950392 | 7 | 12263538 | 8437 | 0.00 | 0.00 | 0.00 | 0.00 | 1.00 |
| CAD | rs1371187 | 2 | 165045101 | 3939 | 0.00 | 0.00 | 0.00 | 0.05 | 0.95 |
| CAD | rs13002621 | 2 | 164915279 | 3898 | 0.00 | 0.00 | 0.00 | 0.05 | 0.95 |
| CAD | rs28582094 | 15 | 38843887 | 5168 | 0.00 | 0.02 | 0.00 | 0.03 | 0.95 |
| CAD | rs2894699 | 7 | 114059156 | 3591 | 0.00 | 0.00 | 0.00 | 0.13 | 0.87 |
| CAD | rs798549 | 7 | 2760750 | 6973 | 0.00 | 0.00 | 0.00 | 0.14 | 0.86 |
| CAD | rs10267593 | 7 | 1937261 | 6164 | 0.00 | 0.00 | 0.00 | 0.14 | 0.86 |
| IS | rs10950392 | 7 | 12263538 | 7982 | 0.00 | 0.03 | 0.00 | 0.02 | 0.95 |

###### **Supplemental Table 11:** SMR and HEIDI analysis of the effect of gene expression in the plaque on major depression

Results for all genes included in the analysis. *SMR*: summary data based Mendelian Randomization; *HEIDI*: heterogeneity in dependent instruments; *Chr*: chromosome; *Beta (SE)*: causal estimate (standard error); *FDR*: False Discovery Rate, *N SNPs*: number of SNPs included in the HEIDI test.

|  |  | SMR | | | HEIDI | |
| --- | --- | --- | --- | --- | --- | --- |
| Gene | **Chr** | **Beta (SE)** | **P-value** | **P-value (FDR)** | **N SNPs** | **P-value** |
| *ADAP1* | 7 | -0.0045 (0.0147) | 0.759 | 0.875 | 12 | 0.543 |
| *BAHD1* | 15 | 0.0133 (0.0268) | 0.620 | 0.845 | 6 | 0.332 |
| *CHAF1A* | 19 | 0.0029 (0.0184) | 0.875 | 0.937 | NA | NA |
| *DISP2* | 15 | -0.0625 (0.0584) | 0.284 | 0.711 | NA | NA |
| *FAM98B* | 15 | 0.0147 (0.0432) | 0.733 | 0.875 | 3 | 0.214 |
| *GNA11* | 19 | -0.0785 (0.0538) | 0.144 | 0.542 | NA | NA |
| *GPR176* | 15 | -0.0106 (0.0196) | 0.588 | 0.845 | NA | NA |
| *KNL1* | 15 | 0.0303 (0.0537) | 0.572 | 0.845 | NA | NA |
| *MDFIC* | 7 | 0.077 (0.0464) | 0.097 | 0.486 | NA | NA |
| *MPND* | 19 | 0.0096 (0.0184) | 0.601 | 0.845 | NA | NA |
| *NDUFA11* | 19 | 0.0274 (0.0396) | 0.489 | 0.845 | NA | NA |
| *PIP5K1C* | 19 | -0.0358 (0.0333) | 0.281 | 0.711 | NA | NA |
| *SUN1* | 7 | -0.0575 (0.0328) | 0.079 | 0.486 | 15 | 0.072 |
| *TLE5* | 19 | 0 (0.0358) | 1.00 | 1.00 | 3 | 0.163 |
| *ZFAND2A* | 7 | 0.0595 (0.0241) | 0.013 | 0.201 | 15 | 0.870 |

###### **Supplemental Table 12:** SMR and HEIDI analysis of the effect of gene expression in the plaque on coronary artery disease

Results for all genes included in the analysis. *SMR*: summary data based Mendelian Randomization; *HEIDI*: heterogeneity in dependent instruments; *Chr*: chromosome; *Beta (SE)*: causal estimate (standard error); *FDR*: False Discovery Rate, *N SNPs*: number of SNPs included in the HEIDI test.

|  |  | SMR | | | HEIDI | |
| --- | --- | --- | --- | --- | --- | --- |
| Gene | **Chr** | **Beta (SE)** | **P-value** | **P-value (FDR)** | **N SNPs** | **P-value** |
| ADAP1 | 7 | -0.0073 (0.0309) | 0.812 | 0.877 | 18 | 0.593 |
| ARRDC5 | 19 | -0.057 (0.0394) | 0.147 | 0.451 | NA | NA |
| BAHD1 | 15 | 0.0576 (0.0551) | 0.296 | 0.504 | 10 | 0.906 |
| CHAF1A | 19 | -0.0058 (0.0379) | 0.877 | 0.877 | NA | NA |
| DISP2 | 15 | 0.331 (0.1458) | 0.0231 | 0.197 | NA | NA |
| FAM98B | 15 | 0.1029 (0.0873) | 0.239 | 0.451 | 3 | 0.645 |
| GNA11 | 19 | -0.0233 (0.0945) | 0.805 | 0.877 | 3 | 0.549 |
| GPR176 | 15 | -0.008 (0.0412) | 0.845 | 0.877 | NA | NA |
| KNL1 | 15 | -0.1312 (0.1072) | 0.221 | 0.451 | NA | NA |
| MDFIC | 7 | 0.1296 (0.0954) | 0.174 | 0.451 | NA | NA |
| MICOS13 | 19 | 0.1543 (0.1024) | 0.132 | 0.451 | 4 | 0.0461 |
| MPND | 19 | 0.0518 (0.0397) | 0.192 | 0.451 | NA | NA |
| NDUFA11 | 19 | 0.104 (0.0875) | 0.234 | 0.451 | NA | NA |
| PIP5K1C | 19 | 0.0603 (0.0668) | 0.367 | 0.568 | NA | NA |
| SUN1 | 7 | -0.0523 (0.0701) | 0.456 | 0.645 | 19 | 0.651 |
| TLE5 | 19 | -0.2093 (0.0913) | 0.0219 | 0.197 | 3 | 0.582 |
| ZFAND2A | 7 | -0.0188 (0.0427) | 0.66 | 0.862 | 20 | 0.137 |

###### **Supplemental Table 13:** SMR and HEIDI analysis of the effect of gene expression in the plaque on stroke

Results for all genes included in the analysis. *SMR*: summary data based Mendelian Randomization; *HEIDI*: heterogeneity in dependent instruments; *Chr*: chromosome; *Beta (SE)*: causal estimate (standard error); *FDR*: False Discovery Rate, *N SNPs*: number of SNPs included in the HEIDI test.

|  |  | SMR | | | HEIDI |  |
| --- | --- | --- | --- | --- | --- | --- |
| Gene | **Chr** | **Beta (SE)** | **P-value** | **P-value (FDR)** | **N SNPs** | **P-value** |
| *ADAP1* | 7 | 0.0557 (0.0437) | 0.202 | 0.821 | 18 | 0.226 |
| *ARRDC5* | 19 | 0.0572 (0.0528) | 0.278 | 0.821 | NA | NA |
| *BAHD1* | 15 | 0.031 (0.0686) | 0.652 | 0.923 | 6 | 0.115 |
| *CHAF1A* | 19 | 0.0262 (0.0518) | 0.613 | 0.923 | NA | NA |
| *DISP2* | 15 | 0.4857 (0.216) | 0.0245 | 0.249 | NA | NA |
| *FAM98B* | 15 | -0.0283 (0.1121) | 0.801 | 0.936 | 3 | 0.23 |
| *GNA11* | 19 | -0.1446 (0.1366) | 0.29 | 0.821 | 3 | 0.563 |
| *GPR176* | 15 | 0 (0.0456) | 1 | 1 | NA | NA |
| *KNL1* | 15 | 0.0215 (0.1428) | 0.881 | 0.936 | NA | NA |
| *MDFIC* | 7 | -0.0197 (0.1183) | 0.868 | 0.936 | NA | NA |
| *MICOS13* | 19 | 0.3331 (0.1528) | 0.0293 | 0.249 | 4 | 0.272 |
| *MPND* | 19 | 0.0417 (0.0537) | 0.437 | 0.888 | NA | NA |
| *NDUFA11* | 19 | 0.0851 (0.1179) | 0.47 | 0.888 | NA | NA |
| *PIP5K1C* | 19 | -0.0598 (0.0942) | 0.526 | 0.894 | NA | NA |
| *SUN1* | 7 | -0.0917 (0.1103) | 0.406 | 0.888 | 16 | 0.597 |
| *TLE5* | 19 | -0.1686 (0.1097) | 0.124 | 0.705 | 3 | 0.258 |
| ZFAND2A | 7 | 0.0117 (0.0572) | 0.837 | 0.936 | 18 | 0.084 |

###### **Supplemental Table 14:** SMR and HEIDI analysis of the effect of gene expression in the plaque on carotid intima-media thickness

Results for all genes included in the analysis. *SMR*: summary data based Mendelian Randomization; *HEIDI*: heterogeneity in dependent instruments; *Chr*: chromosome; *Beta (SE)*: causal estimate (standard error); *FDR*: False Discovery Rate, *N SNPs*: number of SNPs included in the HEIDI test.

|  |  | SMR | | | HEIDI |  |
| --- | --- | --- | --- | --- | --- | --- |
| Gene | **Chr** | **Beta (SE)** | **P-value** | **P-value (FDR)** | **N SNPs** | **P-value** |
| *ADAP1* | 7 | 0.0028 (0.0051) | 0.581 | 0.912 | 18 | 0.916 |
| *ARRDC5* | 19 | 7e-04 (0.0062) | 0.912 | 0.912 | NA | NA |
| *BAHD1* | 15 | -0.0018 (0.008) | 0.824 | 0.912 | 10 | 0.537 |
| *CHAF1A* | 19 | 0.0019 (0.0063) | 0.759 | 0.912 | NA | NA |
| *DISP2* | 15 | -0.02 (0.0256) | 0.436 | 0.868 | NA | NA |
| *FAM98B* | 15 | -0.016 (0.013) | 0.218 | 0.743 | 3 | 0.631 |
| *GNA11* | 19 | -0.0017 (0.0133) | 0.901 | 0.912 | 3 | 0.438 |
| *GPR176* | 15 | -0.0081 (0.0058) | 0.162 | 0.741 | NA | NA |
| *KNL1* | 15 | -0.024 (0.0177) | 0.174 | 0.741 | NA | NA |
| *MDFIC* | 7 | 0.0036 (0.0126) | 0.776 | 0.912 | NA | NA |
| *MICOS13* | 19 | 0.0287 (0.0161) | 0.0744 | 0.632 | 4 | 0.131 |
| *MPND* | 19 | 0.0062 (0.0064) | 0.331 | 0.805 | NA | NA |
| *NDUFA11* | 19 | 0.0158 (0.0143) | 0.27 | 0.766 | NA | NA |
| *PIP5K1C* | 19 | -0.006 (0.012) | 0.619 | 0.912 | NA | NA |
| *SUN1* | 7 | -0.0077 (0.0105) | 0.459 | 0.868 | 18 | 0.4 |
| *TLE5* | 19 | -0.0252 (0.0136) | 0.0633 | 0.632 | 3 | 0.71 |
| *ZFAND2A* | 7 | -0.0016 (0.0063) | 0.803 | 0.912 | 19 | 0.209 |

### Supplemental Figures

###### **Supplemental Figures 1 to 10:** Mirror plots for top results of colocalization analysis (posterior probability for shared causal variant > 0.8).

Panels from top to bottom: SNP associations with major depression, SNP associations with the respective atherosclerotic disease, basic gene annotation. Plots were generated using the R package RACER (https://github.com/oliviasabik/RACER)^27^. ALLSTROKE: any stroke; IS: ischemic stroke; CAD: coronary artery disease; LD_BIN: levels of linkage disequilibrium with lead SNP.

###### **Supplemental Figure 11 to 22.** GTEx multi-tissue plots for lead SNPs from colocalizing regions.

GTEx multi-tissue plots obtained from the GTEx-Portal (<https://www.gtexportal.org/home/>) displaying how a specific eQTL influences expression of a gene across human tissues. Title: Gene (ENSEMBL Gene ID/Gene Symbol), eQTL ID and p-value of association in meta-analysis of tissues. NES: normalized effect size; m-value: parameter for specificity of expression from 0 (low) to 1 (high); CI: confidence interval.

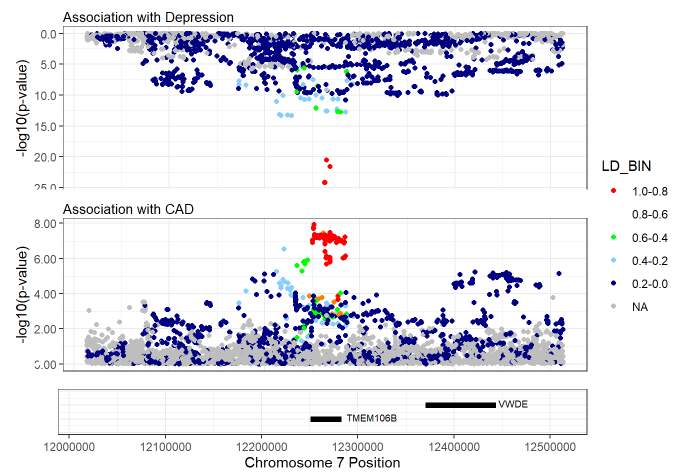

###### **Supplemental Figure 1:** Mirror plot of depression and CAD at rs10950392 on chromosome 7.

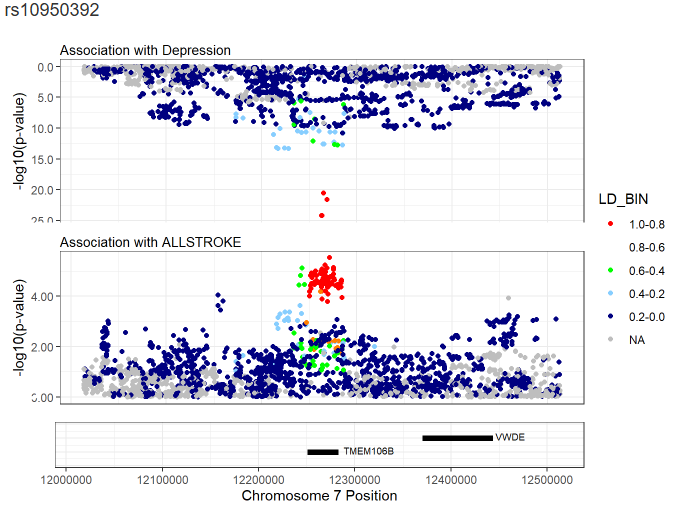

###### **Supplemental Figure 2:** Mirror plot of depression and ALLSTROKE at rs10950392 on chromosome 7.

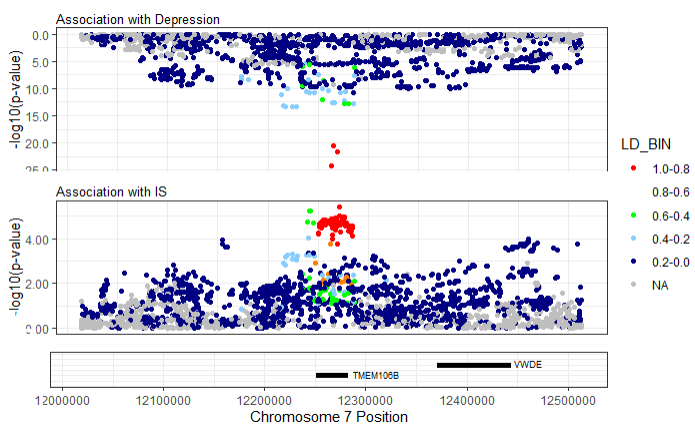

###### **Supplemental Figure 3:** Mirror plot of depression and IS at rs10950392 on chromosome 7.

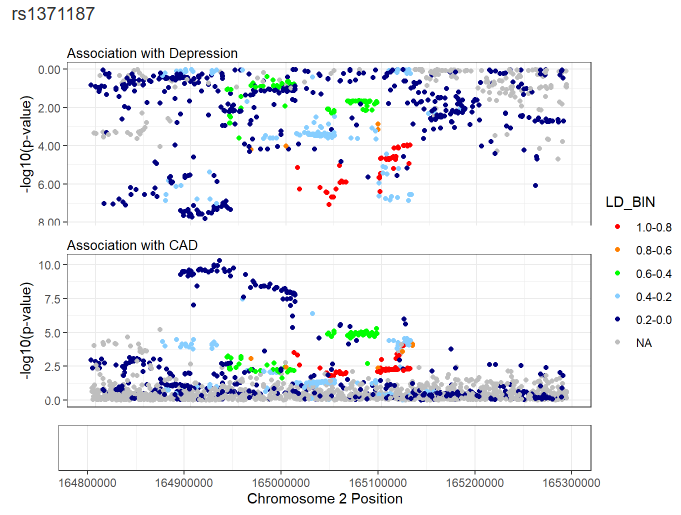

###### **Supplemental Figure 4:** Mirror plot of depression and CAD at rs1371187 on chromosome 2.

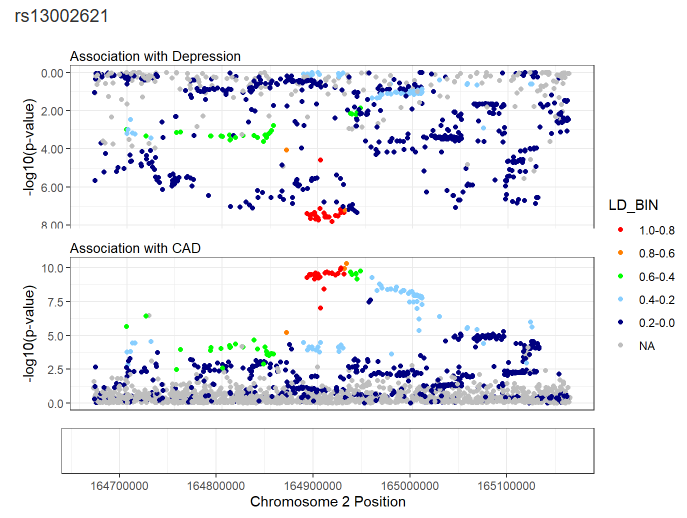

###### **Supplemental Figure 5:** Mirror plot of depression and CAD at rs13002621.

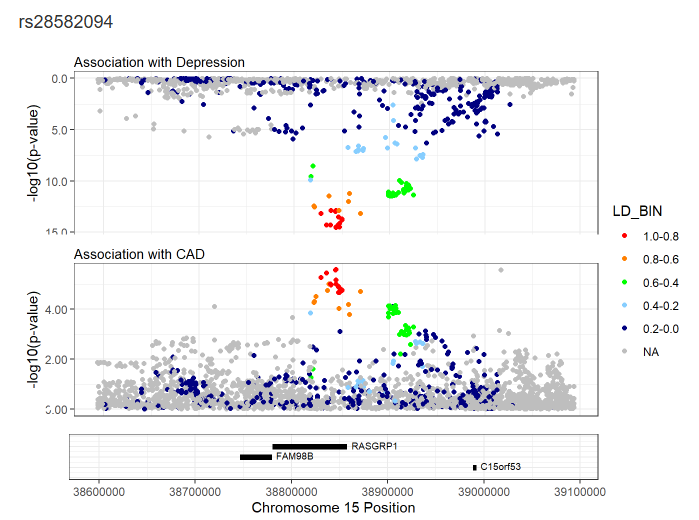

###### **Supplemental Figure 6:** Mirror plot of depression and CAD at rs28582094 on chromosome 15.

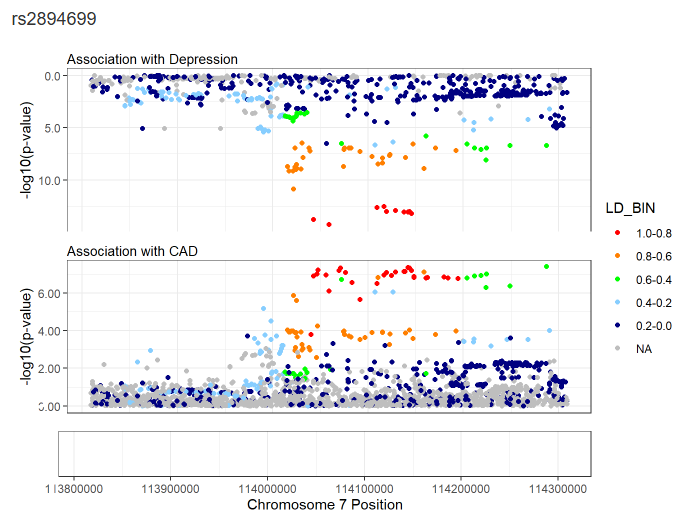

###### **Supplemental Figure 7:** Mirror plot of depression and CAD at rs2894699 on chromosome 7.

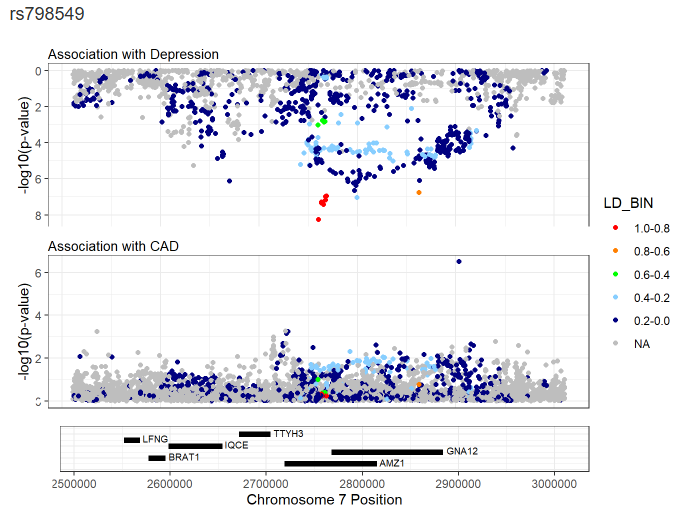

###### **Supplemental Figure 8:** Mirror plot of depression and CAD at rs798549 on chromosome 7.

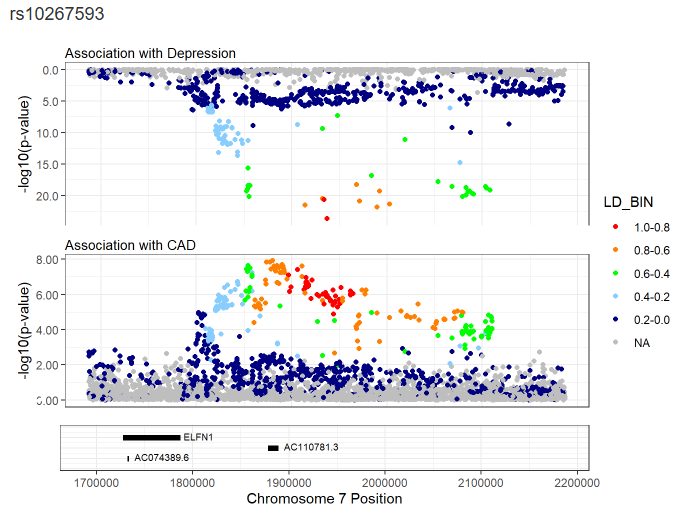

###### **Supplemental Figure 9:** Mirror plot of depression and CAD at rs10267593 on chromosome 7.

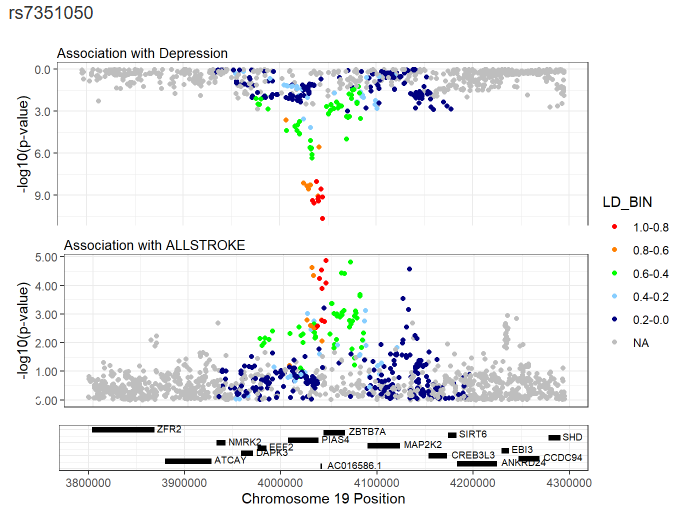

###### **Supplemental Figure 10:** Mirror plot of depression and ALLSTROKE at rs7351050 on chromosome 19.

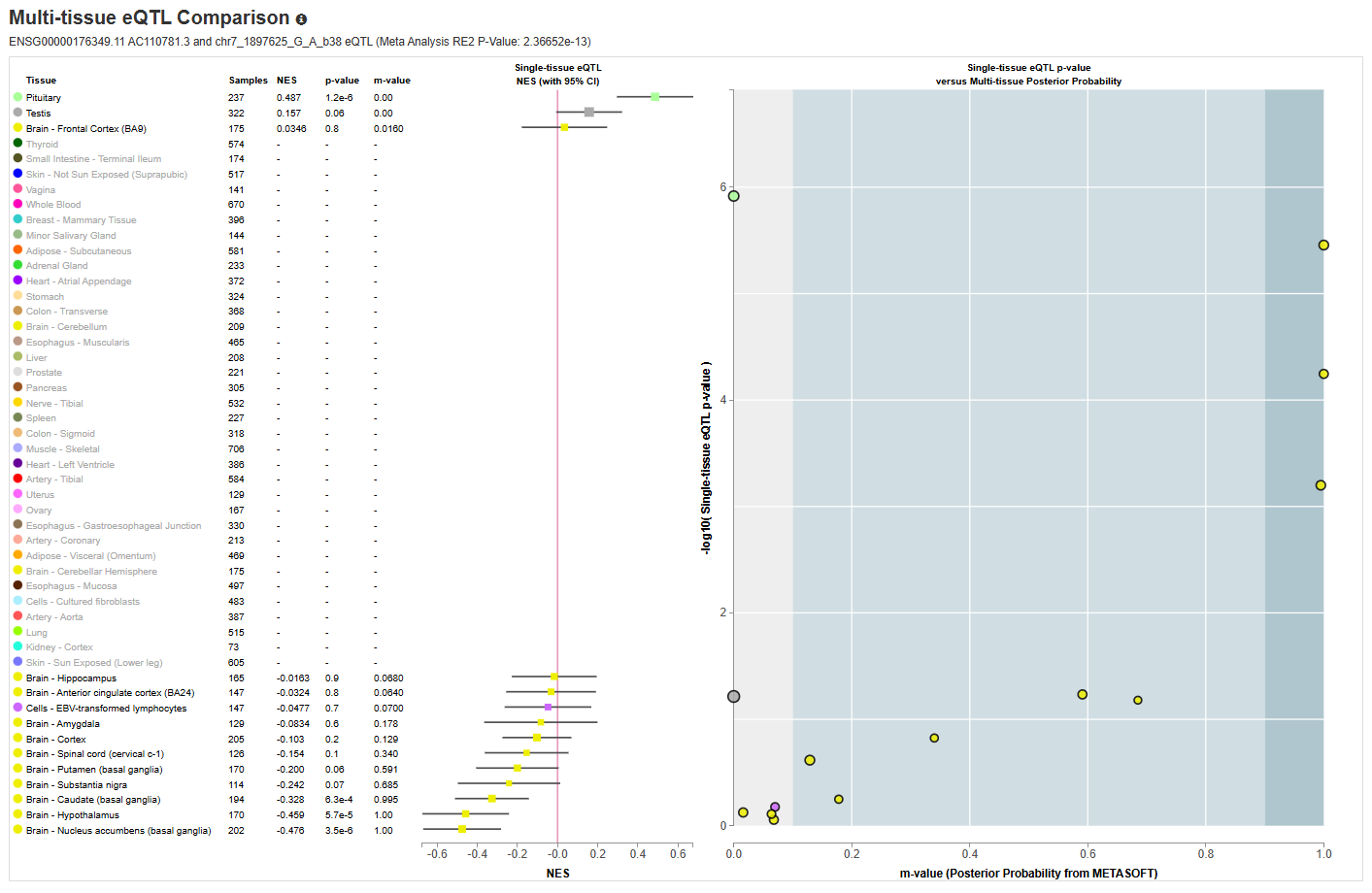

###### **Supplemental Figure 11:** GTEx multi-tissue plot for AC110781.3.

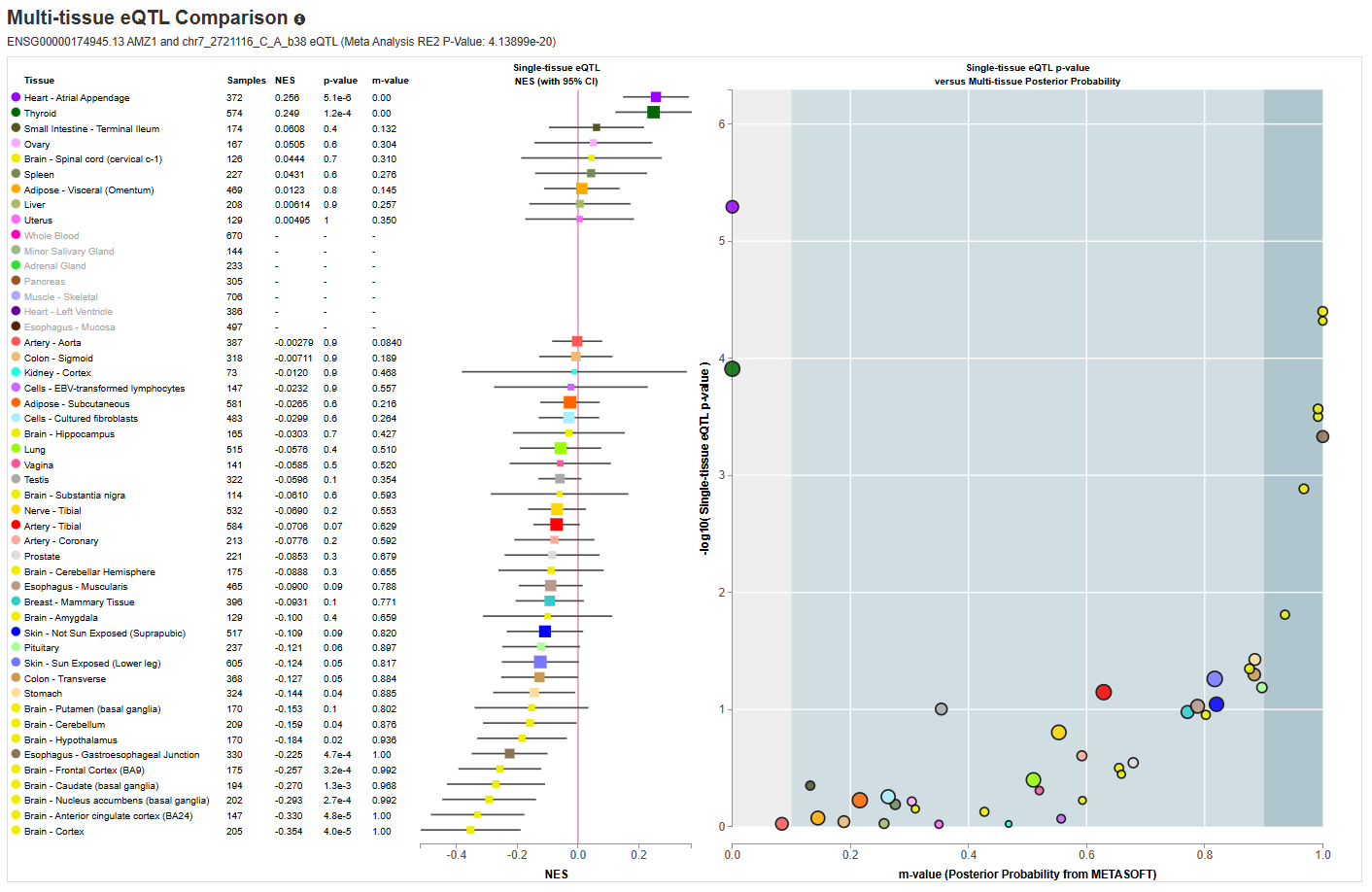

###### **Supplemental Figure 12:** GTEx multi-tissue plot for AMZ1.

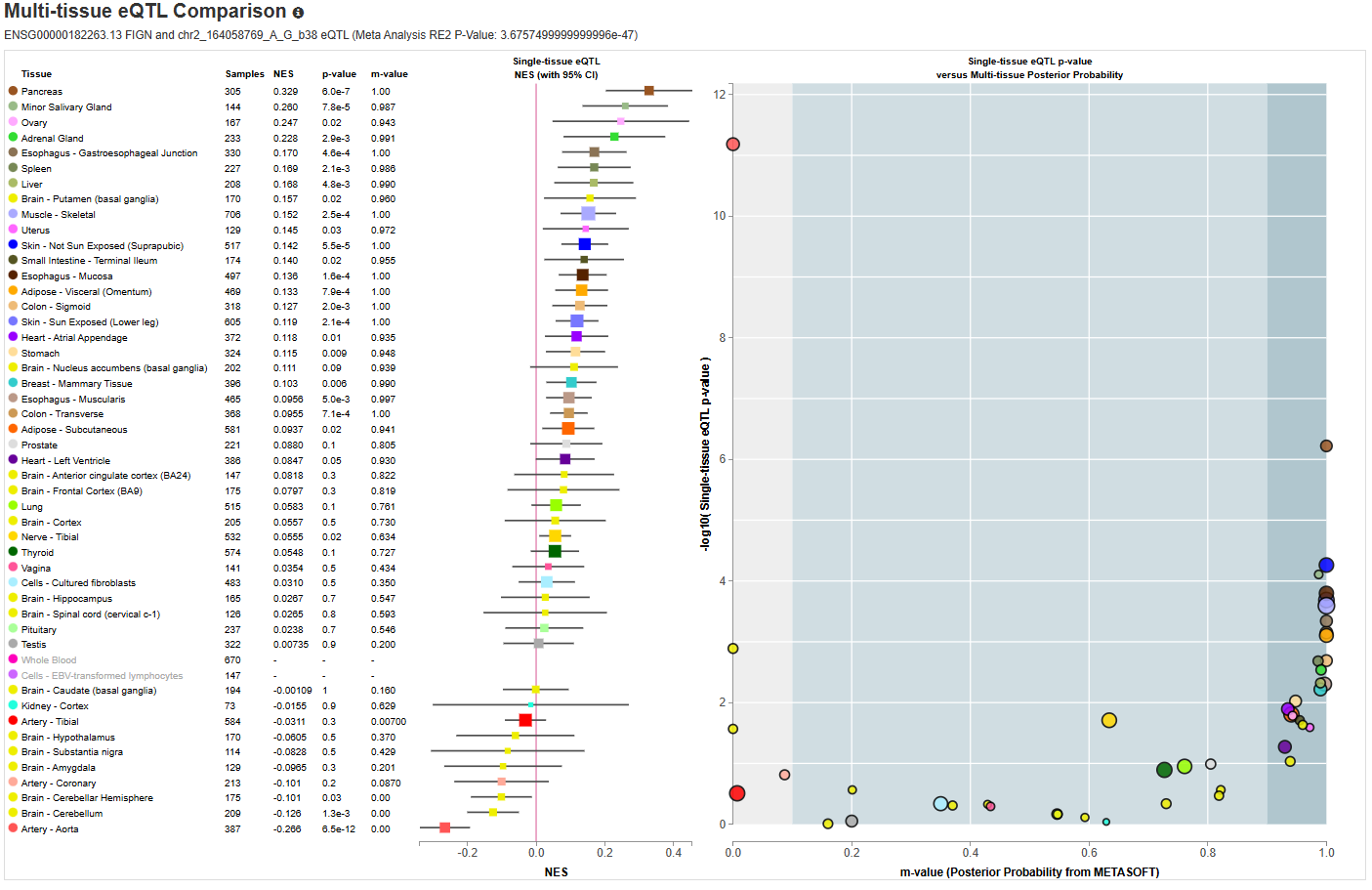

###### **Supplemental Figure 13:** GTEx multi-tissue plot for FIGN.

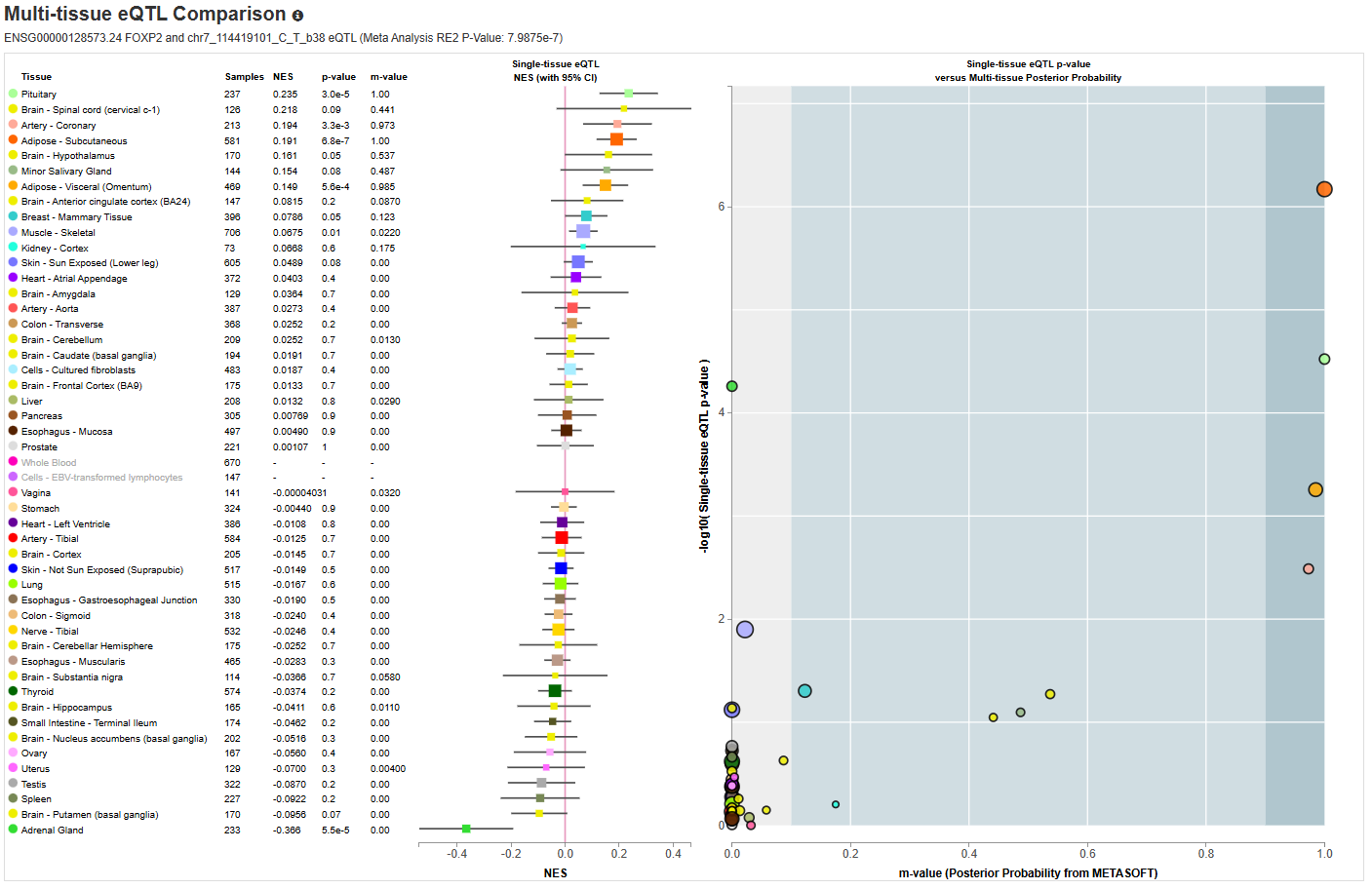

###### **Supplemental Figure 14**: GTEx multi-tissue plot for FOXP2.

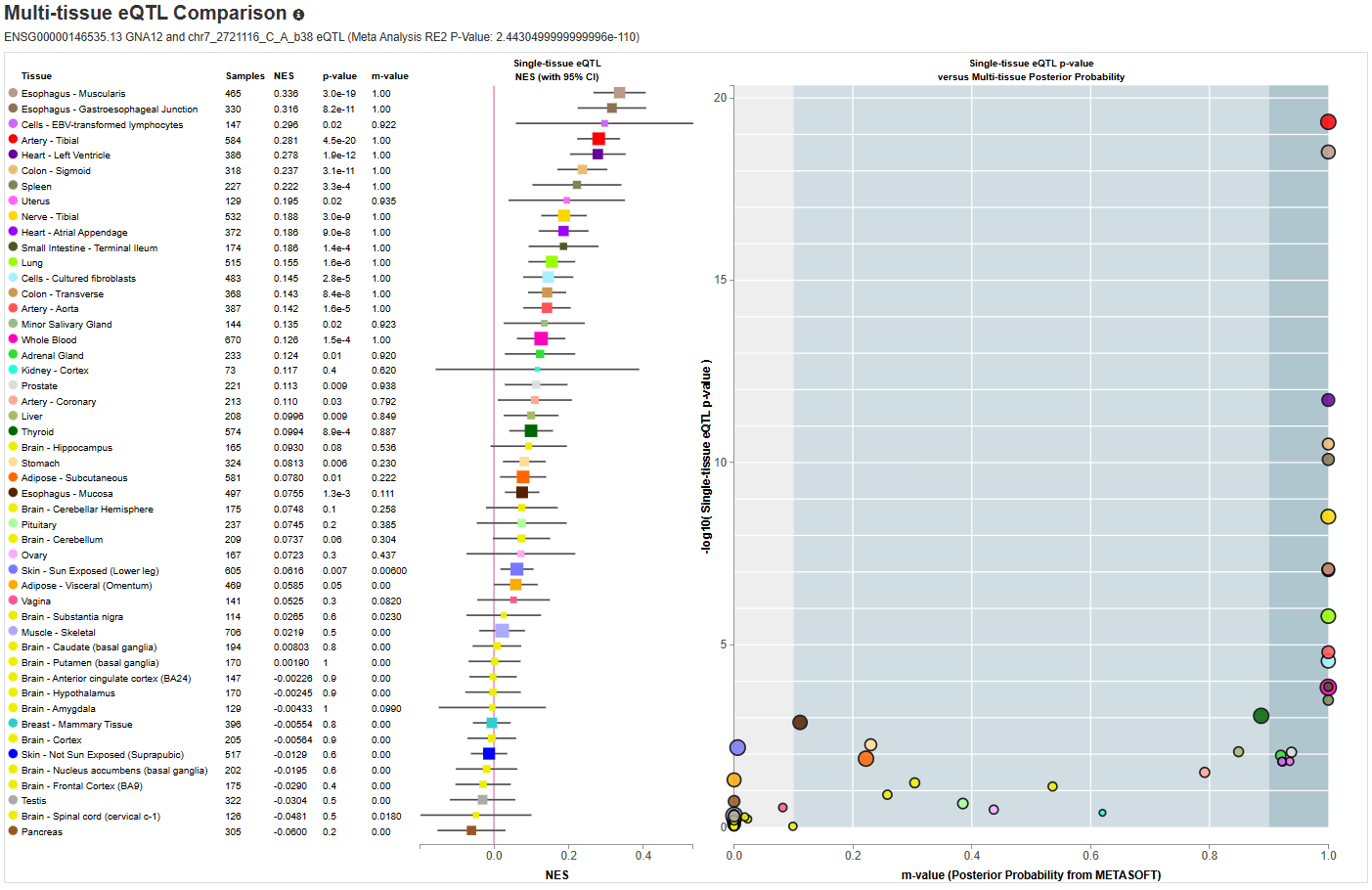

###### **Supplemental Figure 15:** GTEx multi-tissue plot for GNA12.

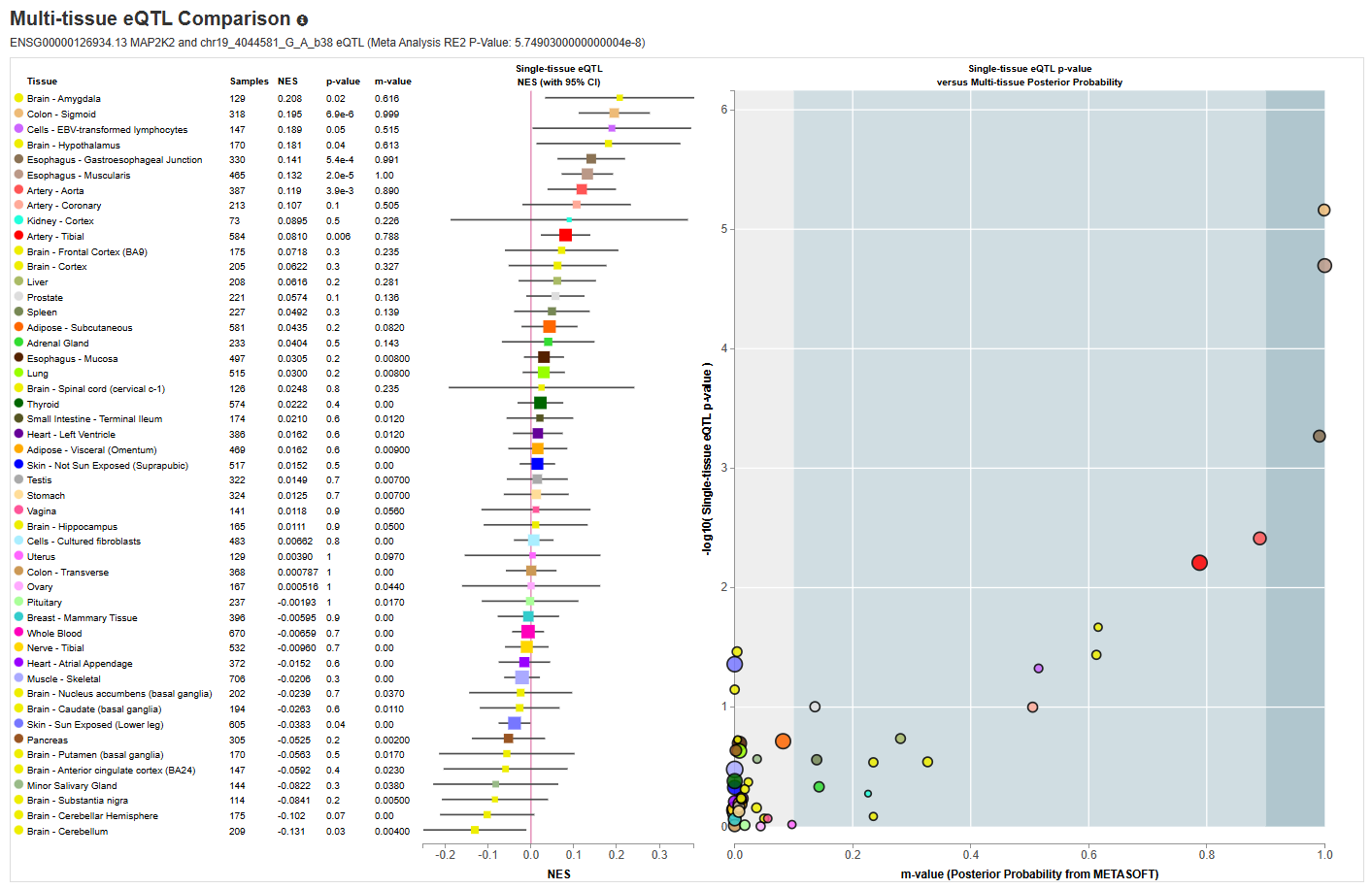

###### **Supplemental Figure 16:** GTEx multi-tissue plot for MAP2K2.

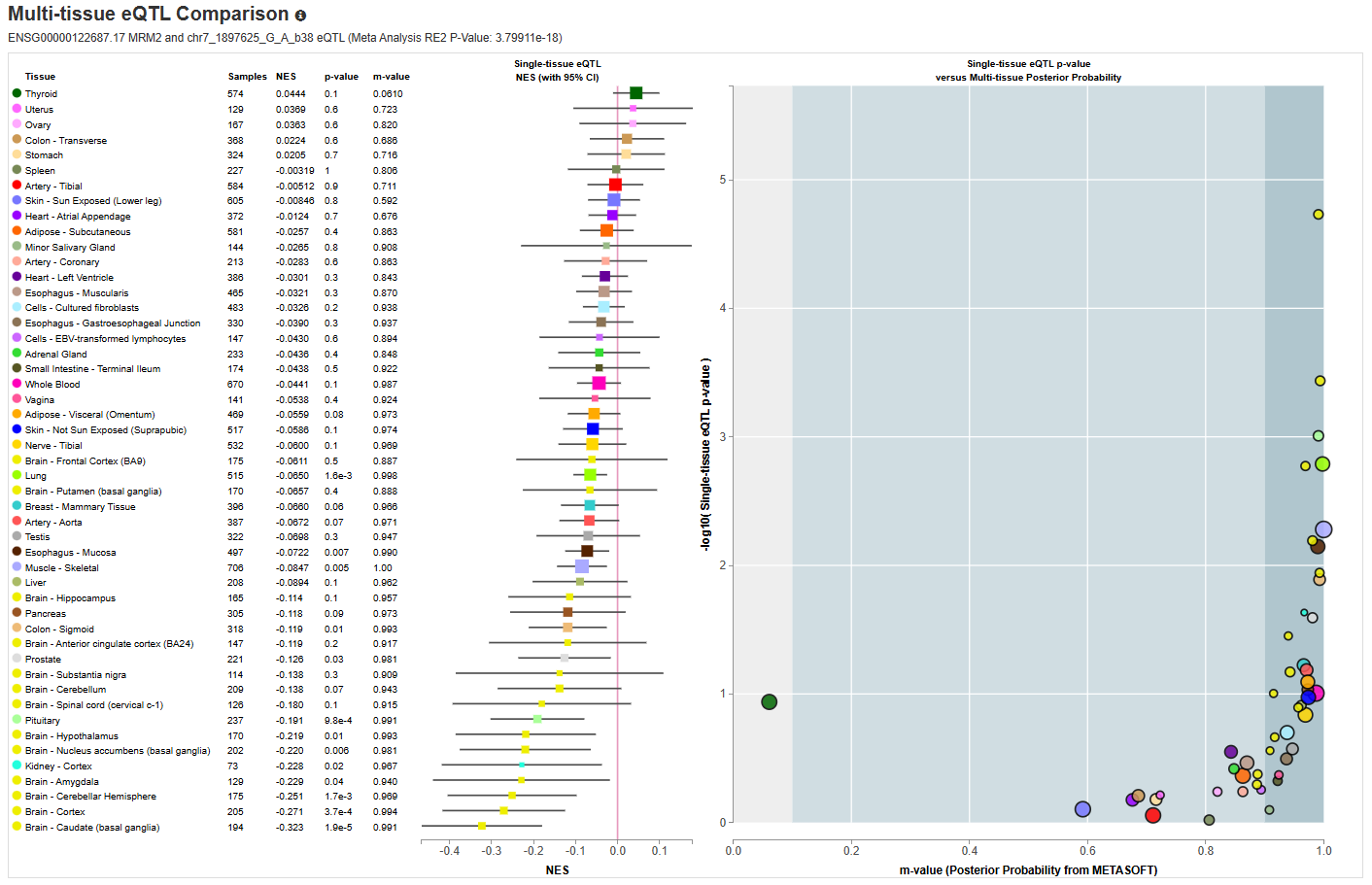

###### **Supplemental Figure 17:** GTEx multi-tissue plot for MRM2.

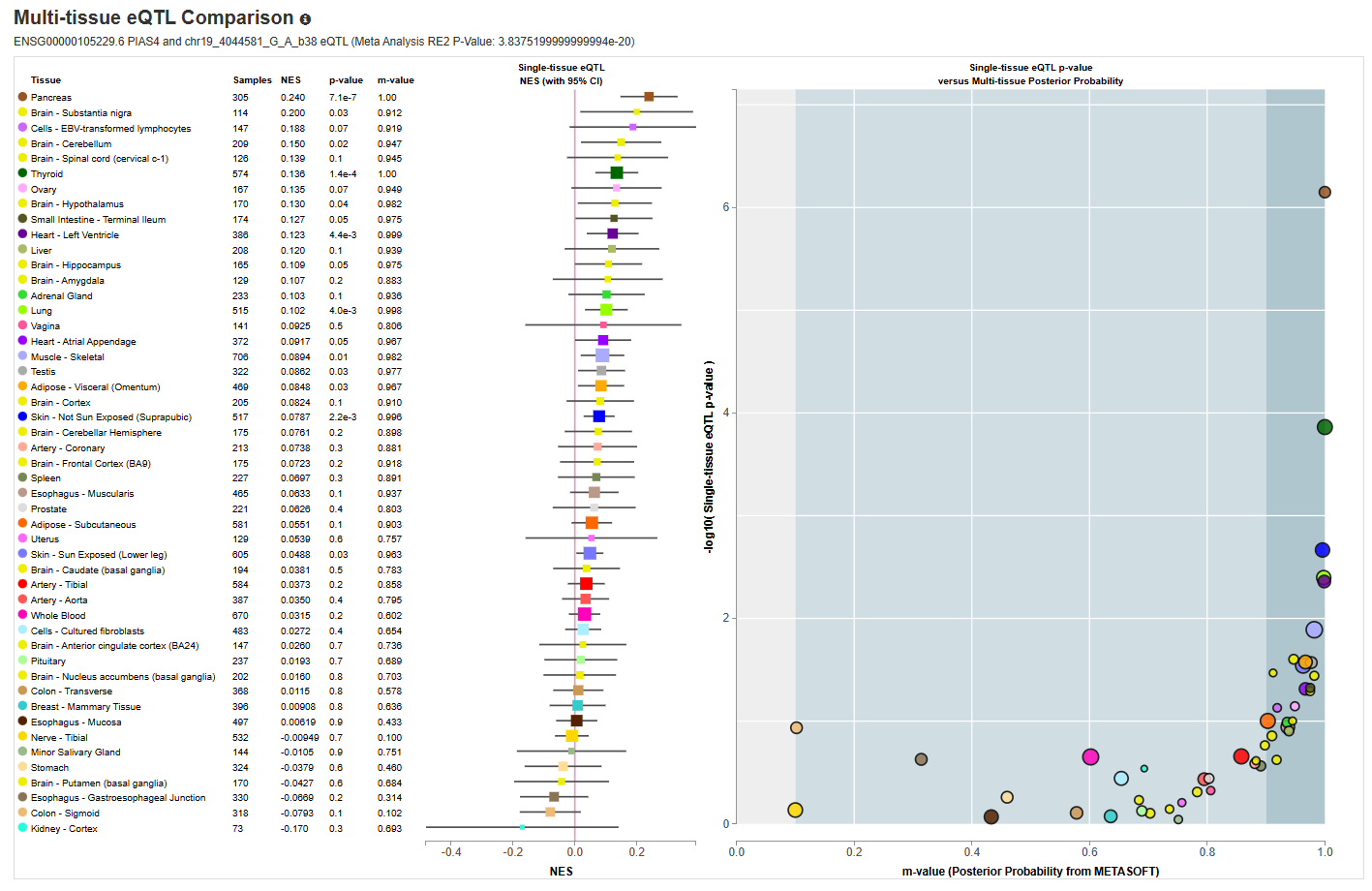

###### **Supplemental Figure 18:** GTEx multi-tissue plot for PIAS4.

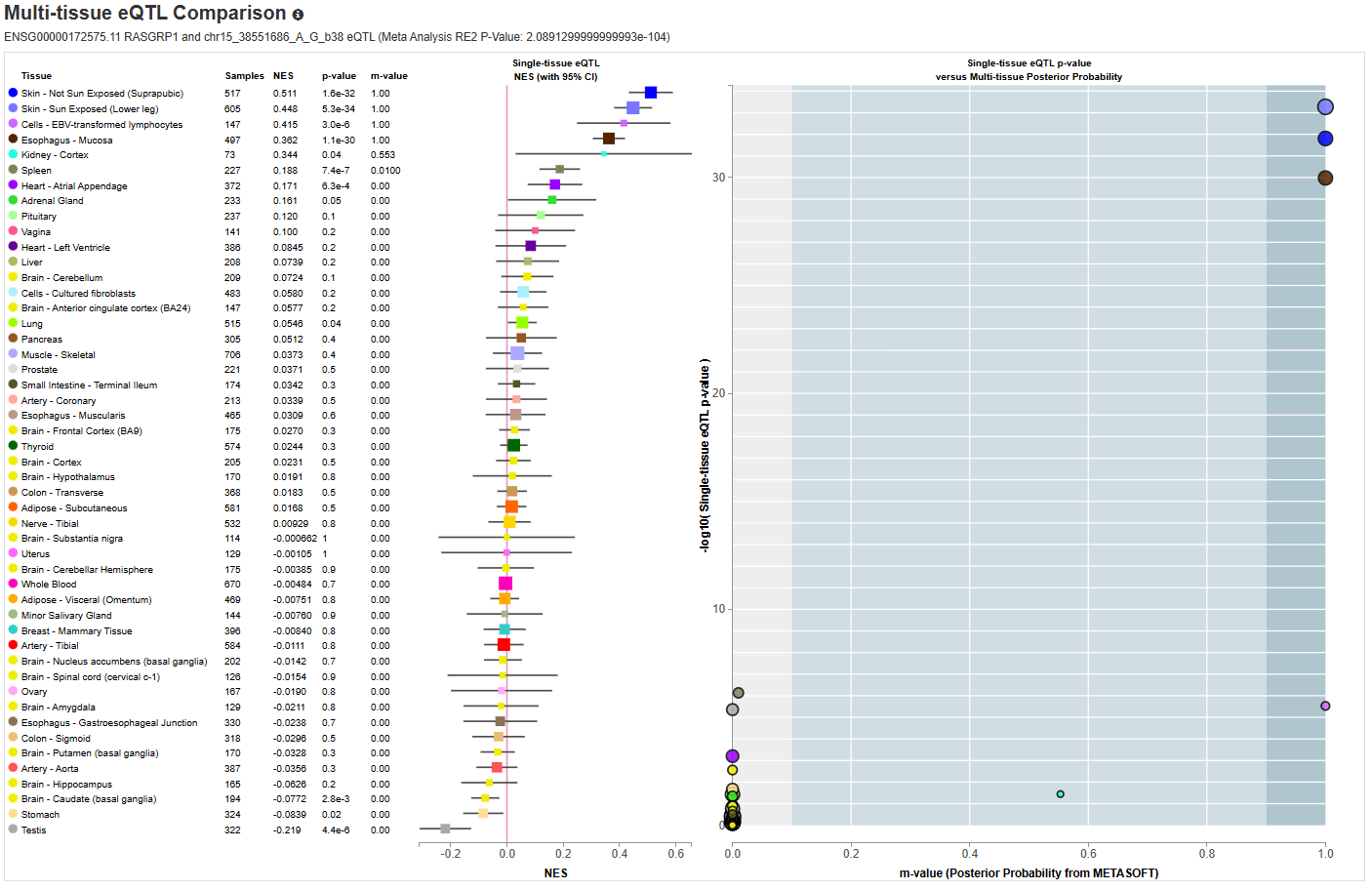

###### **Supplemental Figure 19:** GTEx multi-tissue plot for RASGRP1.

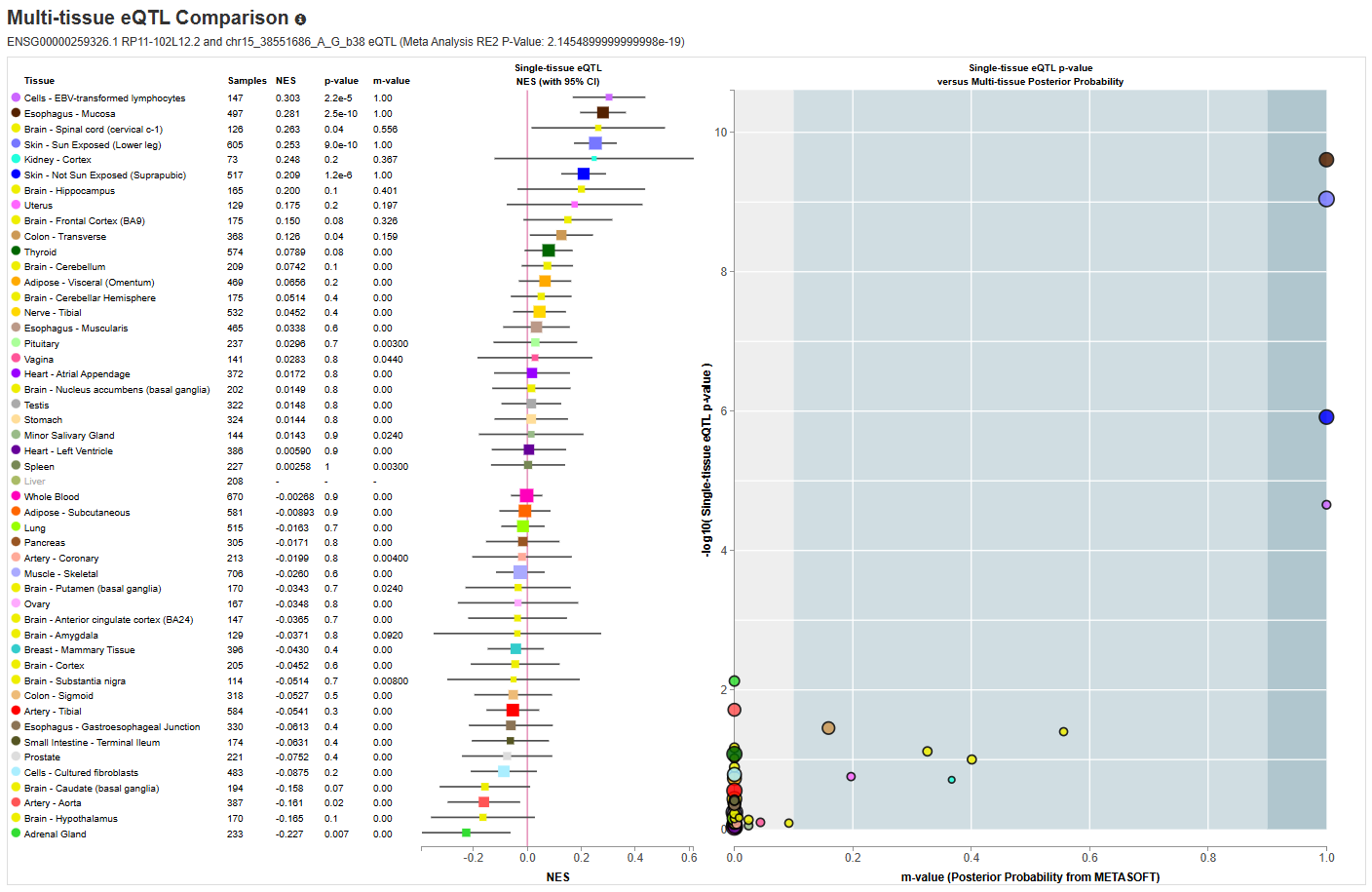

###### **Supplemental Figure 20:** GTEx multi-tissue plot for RP11-102L12.2.

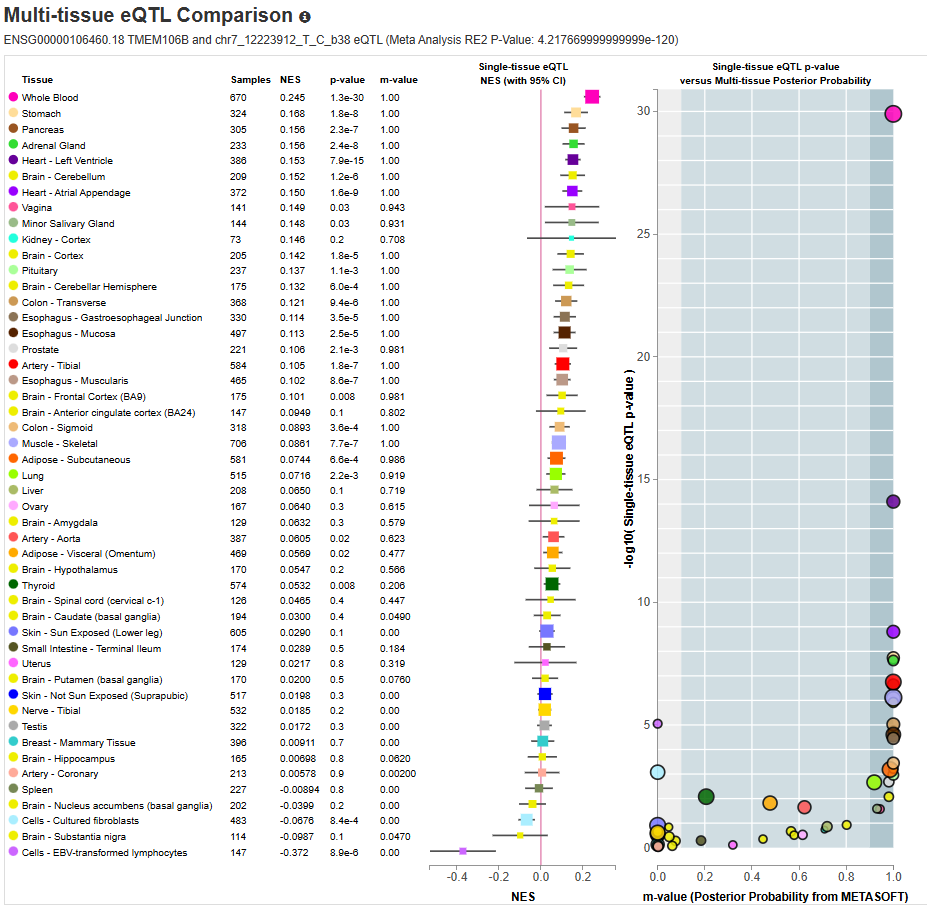

###### **Supplemental Figure 21:** GTEx multi-tissue plot for TMEM106B.

Expression of *TMEM106B* across tissues was found to be significantly associated with lead SNP rs10950392. Significant effects and high tissue specificity were found for – amongst others – whole blood, adrenal gland, left ventricle, atrial appendage, tibial artery, multiple brain parts (cortex, cerebellum, cerebellar hemisphere), pituitary.

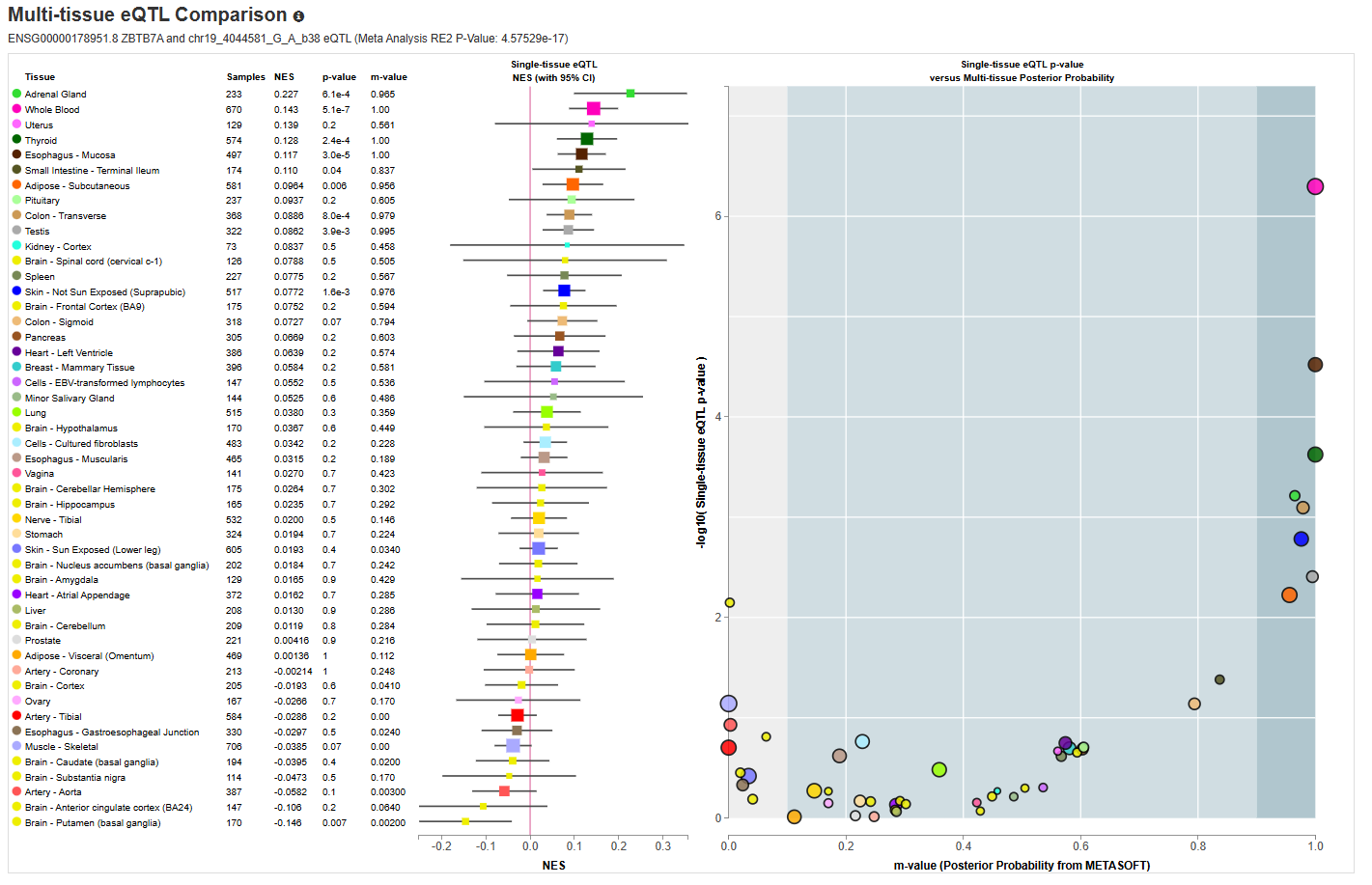

###### **Supplemental Figure 22:** GTEx multi-tissue plot for ZBTB7A.

Expression of *ZBTB7A* across tissues was found to be significantly associated with lead SNP rs7351050. Significant effects and high tissue specificity were found for whole blood and thyroid.

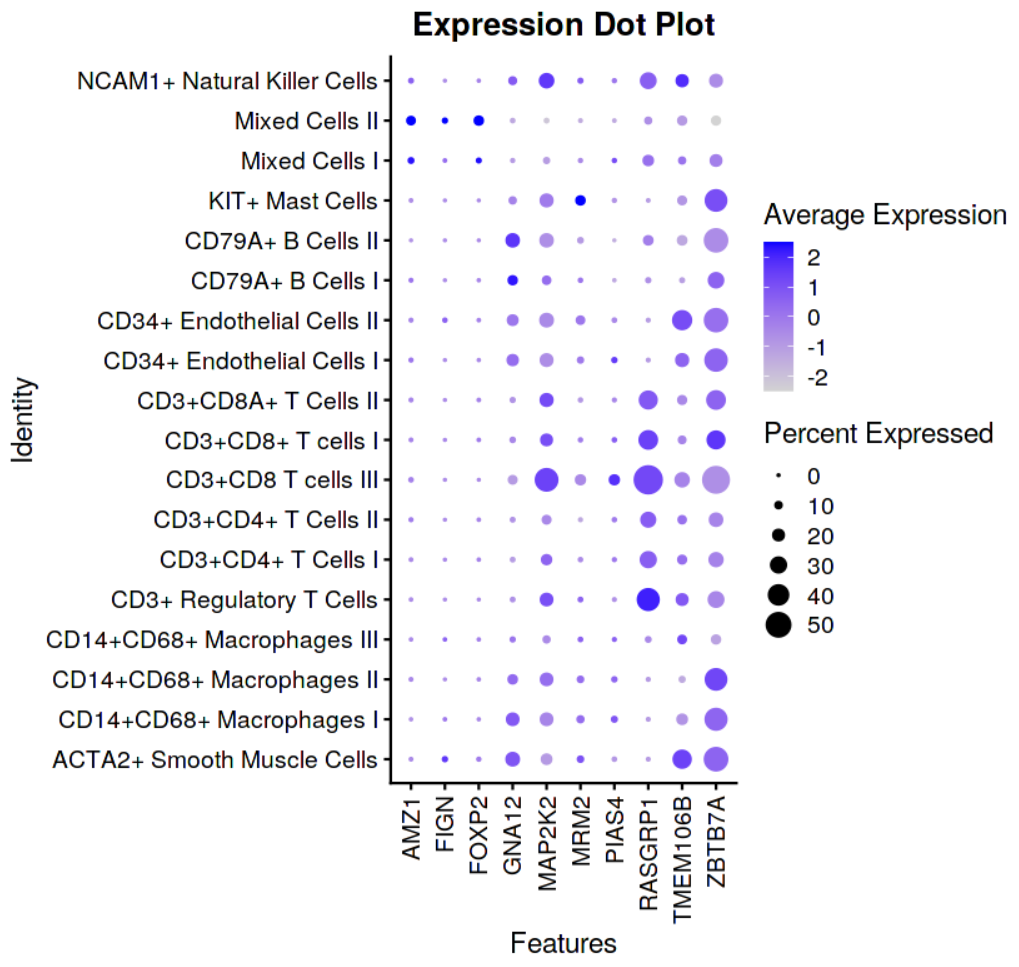

###### **Supplemental Figure 23:** Expression dot plot for GTEx lookup in PlaqView (<https://www.plaqview.com>)^28^.
